# Arts and Cultural Engagement and Multidimensional Well-being in Later Life

**DOI:** 10.64898/2026.06.02.26354582

**Authors:** Taiji Noguchi, Erhua Shang, Takahiro Hayashi

**Author notes:** Corresponding author: Taiji Noguchi, PhD, Department of Community Health and Preventive Medicine, Hamamatsu University School of Medicine, 1-20-1 Handayama, Chuoku, Hamamatsu, Shizuoka 431-3192, Japan.

## Abstract

**Background and Objectives:** Arts and cultural engagement may contribute to well-being in later life. However, evidence from longitudinal studies from Asia, including Japan, remains limited. This study examined the association of arts and cultural engagement with subsequent multidimensional well-being among older adults in Japan, one of the fastest-aging countries.

**Research Design and Methods:** This longitudinal study used panel data from 354 individuals aged 60 and older (mean age 74.0 years; 78.6% women) who completed self-administered questionnaires by mail between 2022 and 2024. The PERMA-Profiler was used to assess five multifaceted aspects of psychological well-being: positive emotion, engagement, relationships, meaning, and accomplishment. Frequencies of arts and cultural engagement at baseline were measured for active (e.g., activities by individuals and participation in groups, such as music and painting) and receptive (e.g., visiting museums, galleries, and theaters) forms.

**Results:** Multivariable linear regression analysis, adjusted for the covariates including baseline PERMA scores, showed that higher frequencies of active engagement were positively associated with higher PERMA scores for all domains. Higher frequencies of receptive engagement were associated with the domains of positive emotion, meaning, and accomplishment, but not clearly associated with engagement and relationships. Restricted cubic spline analyses suggested clearer positive frequency-response patterns for active engagement than for receptive engagement.

**Discussion and Implications:** Arts and cultural engagement, in both active and receptive forms, was associated with multiple aspects of subsequent well-being in later life. These findings suggest the importance of ensuring access to arts and cultural opportunities for older adults to create, participate, and connect.

## Introduction

The number of older adults is increasing more rapidly than any other age group globally. According to the United Nations, the number of those aged 65 or older will more than double, from 761 million in 2021 to 1.6 billion in 2050 (United Nations Department of Economic and Social Affairs, 2023). Japan has one of the most rapidly aging populations worldwide, with more than one in four people aged 65 years and over (Cabinet Office Government of Japan, 2024). In old age, individuals face declines in subjective and psychological well-being due to commonly experienced events such as separation from partners and friends, their own physical and psychological deterioration, living alone, reduced social interaction, low income, and age-related worsening health (National Institute for Health and Care Excellence, 2016). Therefore, it is necessary to investigate sustainable ways to ensure well-being in later life.

Well-being is a multidimensional construct, although its conceptualization and definition remain under active discussion. It is not only limited to the absence of negative psychological states, such as depression, anxiety, or distress, but also includes positive dimensions of mental health, such as positive emotions, meaningful life, social connectedness, and a sense of achievement (Dodge, Daly, Huyton, & Sanders, 2012). Meanwhile, previous theories have distinguished hedonic well-being, which refers to pleasure and positive affect, from eudaimonic well-being, which refers to meaning, purpose, and personal growth (Ryan & Deci, 2001; Steptoe, Deaton, & Stone, 2015). While many well-being models include a composite of hedonic and eudaimonic, positive and negative dimensions, Seligman conceptualized well-being more holistically as human flourishing, combining multiple hedonic and eudaimonic dimensions (Seligman, 2011). This model, which is referred to as PERMA, comprises five domains: including positive emotion (hedonic feelings of happiness, pleasure, and comfort), engagement (deep psychological connection to particular activities or organizations), relationships (feelings of interaction with community, links with loved ones, and satisfaction with social networks), meaning (sense of purpose in one’s life direction), and accomplishment (sense of progress toward one’s goals and achievement of superior results). This framework provides a useful model for capturing the multidimensional aspects of human flourishing and is well adapted to the life-course approach to aging (Bartholomaeus, Van Agteren, Iasiello, Jarden, & Kelly, 2019; Sturmberg, 2013; Wister et al., 2016).

There is a growing body of evidence that arts and cultural engagement contribute to people’s health and well-being (Fancourt & Finn, 2019; Liu et al., 2026; Sonke et al., 2025), and it is recognized as a positive behavior that supports longer lives that are better lived (All-Party Parliamentary Group on Arts Health and Wellbeing, 2017; Fancourt et al., 2026; Mak et al., 2026; Rodriguez et al., 2024). The arts are commonly defined as activities which are valued in their own right, provide imaginative experiences, and provoke emotional response (Adajian, 2022; Mizuno & Xu, 2022), such as music, creative writing, crafts, visiting museums and galleries, and attending arts-based community events (Davies et al., 2012; J. Sonke et al., 2023). Arts and cultural engagement is often divided into those that are more active (i.e., requiring the creation of or participation in the arts) and those that are more receptive (i.e., involving arts that have been created and are now experienced by an audience) (Davies, et al., 2012; Holt, Tischler, Vougioukalou, & Corvo, 2025). In both forms, engaging in arts and culture can be considered an activity with multimodal elements containing a variety of “active ingredients” that are beneficial to mental health in later life, including cognitive stimulation, social interaction, multi-sensory engagement, and opportunities for creativity, imagination, and meaning-making (Warran, Burton, & Fancourt, 2022).

Mechanistically, these ingredients may activate psychological (e.g., improving self-esteem and buffering stress), social (e.g., reducing loneliness and enhancing communication), physical (e.g., reducing stress hormones and inflammation), and behavioral (e.g., encouraging health-promoting behaviors) pathways (Fancourt, Aughterson, Finn, Walker, & Steptoe, 2021; Fancourt & Finn, 2019). These effects are unlikely to be uniform, and the outcomes may vary depending on the forms of engagement and individual contexts, including socioeconomic backgrounds.

Evidence from intervention studies has been supported and extended by observational epidemiological studies, which suggest that arts and cultural engagement in daily life can enhance the well-being of older adults. Several previous studies from Western countries have shown the beneficial effects of engaging in arts and cultural activities on various aspects of well-being. For instance, longitudinal studies found that frequent arts and cultural engagement was associated with higher levels of life satisfaction, happiness, worthwhile life, and mental well-being among older adults (Davies, Budgeon, Murray, Hunter, & Knuiman, 2023; Finn, Bone, Fancourt, Warran, & Mak, 2025; Tymoszuk, Perkins, Spiro, Williamon, & Fancourt, 2020). Arts also indicate a reduction in the development of depression and a decrease in anxiety and loneliness in later life (Bone et al., 2022; Fancourt & Tymoszuk, 2019; Finn, et al., 2025). These findings suggest that art and cultural engagement can contribute to multiple aspects across hedonic and eudemonic well-being.

However, most evidence comes from Western countries, and it remains unclear whether a beneficial association between arts and cultural engagement and well-being among older adults can also be observed in East Asia, including Japan. Because East Asia is aging at a rapidly accelerating rate, enhancing the well-being of older adults in this region is a global issue. Additionally, there are considerable differences in social attitudes toward arts and culture, including national resources and educational opportunities, across Western and East Asian countries (Agency for Cultural Affairs, 2021; Naoe, 2003). These cultural contextual differences highlight the necessity of examining people’s experiences of well-being through the arts across countries and cultures. Furthermore, previous studies on arts and well-being have focused on either active or receptive engagement, and evidence remains insufficient on the simultaneous evaluation of both forms. Given the diverse forms of arts, it is important to investigate both engagements in view of policy-making to offer arts and culture for older adults.

Therefore, this study aimed to examine the association between arts and cultural engagement and subsequent well-being among older adults in Japan, which is one of the fastest-aging countries, focusing on both active and receptive forms. We investigated the prospective association with multidimensional well-being, using longitudinal data.

## Methods

### Study participants

This longitudinal study used panel data from two waves conducted in 2022 and 2024. We recruited community-dwelling individuals at public facilities, such as community and cultural centers, in Aichi Prefecture, which is located in the central part of Japan. Details about the baseline survey are provided elsewhere (Noguchi & Shang, 2023). Figure 1 illustrates the sample selection flow. A questionnaire-based survey was conducted between July and August 2022. A total of 1,000 individuals were distributed questionnaires at the venue, and 648 people responded by mail (response rate = 64.8%). Of the respondents, those who did not agree to participate in the study (n = 38), those without information on age and/or sex (n = 5), and those aged under 60 years (n = 83) were excluded. Subsequently, 522 participants were followed two years later, from July to August 2024, by a mailed questionnaire, and 355 responded (response rate = 68.0%). Of them, one individual was excluded because of their disapproval of participating in the study. Thus, a total of 354 individuals were included in the final analysis.

**Figure 1.**
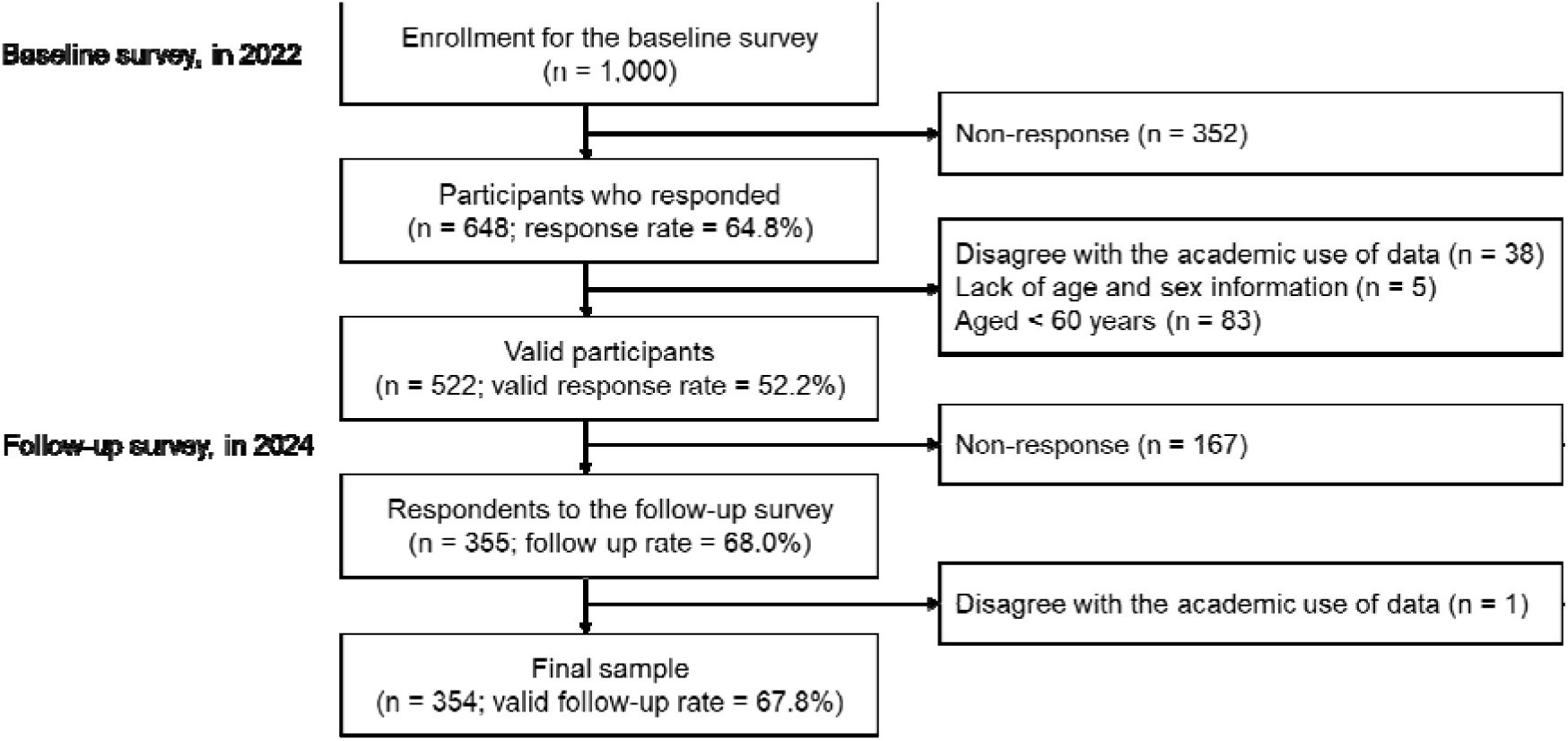
Sample selection flow

This study was approved by the Ethics Committee of the National Center for Geriatrics and Gerontology (No. 1588 and 1801) and Aichi Toho University (No. 202118). The questionnaire was accompanied by a study explanation, and the participants returned a complete questionnaire and written consent to participate in the study. All the procedures were performed in accordance with the Declaration of Helsinki.

### Psychological well-being

In both baseline and follow-up surveys, participants completed the PERMA-Profiler questionnaires (Butler & Kern, 2016), the Japanese version (Watanabe et al., 2018). This instrument is a multidimensional well-being scale, comprising 15 items based on the five domains of the PERMA model by Seligman (Seligman, 2018; Seligman, 2011): (i) positive emotion (3 items related to hedonic feelings, such as happiness, pleasure, and comfort); (ii) engagement (3 items related to deep psychological connection with particular activities, organizations or causes, such as being interested, engaged, and absorbed); (iii) relationships (3 items related to feelings of integration with community members, being cared for loved ones and being satisfied with one’s social network); (iv) meaning (3 items related to a sense of purpose and direction in life, and feeling connected to something larger than the self); and (v) accomplishment (3 items relate to a sense of progressing toward one’s goals and achieving superior results). Additionally, we assessed the filler items of the PERMA-Profiler for four domains, with overall well-being, physical health (3 items), negative emotion (3 items), and loneliness (1 item); a domain of overall well-being was calculated as the mean of the 15 PERMA-Profiler items plus a single item on sense of happiness. Each item was rated on a 0–10 Likert scale (0 = extremely low and 10 = extremely high). We calculated the composite scores for each domain by taking the mean of the items in that domain, with scores ranging from 0 to 10, and higher scores indicating higher well-being.

### Arts and cultural engagement

Based on previous studies (Finn, et al., 2025; Noguchi et al., 2022; Noguchi & Shang, 2023), active and receptive arts and cultural engagement were measured at the baseline. Active engagement was defined as the following nine items: musical performance, singing, dancing, painting, handi-crafting, photography, and three traditional Japanese cultural activities (haiku/tanka/senryu [poetry], kado/sado [flower arrangement/tea ceremony], and shodo [calligraphy]). Receptive engagement was defined as the following four items: going to a film at a cinema, visiting museums and arts galleries, going to theaters, and going to concerts and opera. The participants responded to the frequency of each activity (possible answers: “never,” “once a year,” “a few times a year,” “once a month,” “a few times a month,” or “more than once a week”). The frequencies were summed up for each active and receptive engagement, and divided into three groups: none, yearly to monthly, and monthly or more.

### Covariates

According to previous research (Noguchi & Shang, 2023), the covariates included age, gender, living arrangement, marital status, educational attainment, subjective economic status, employment status, number of illnesses, self-reported health, instrumental activities of daily living (IADL) performance, motor function, subjective cognitive function, drinking, and smoking at baseline. Age (years) was categorized as 60–69, 70–79, and ≥ 80. Living arrangement was dichotomized as living alone and living together. Marital status was dichotomized as not married (never married, divorced, or separated) and married. Educational attainment (years) was categorized as < 10, 10–12, and ≥ 13. Subjective economic status was categorized as low, middle, and high. Employment status was dichotomized as not employed and employed. The number of illnesses from the list of 17, including cancer, stroke, heart disease, diabetes, digestive diseases, dementia, and depression disorders, was categorized as none, one, two, and three or more. Self-reported health was dichotomized as poor and good. IADL performance was assessed using a 5-point subscale of the Tokyo Metropolitan Institute of Gerontology Index of Competence (Koyano, Shibata, Nakazato, Haga, & Suyama, 1991), and the participants were dichotomized as “with difficulty” (when difficulty in performing at least one item was reported) and “without difficulty.” Motor function was assessed using a 5-point subscale of the Kihon checklist (KCL) and dichotomized as not impaired (< 3 points) and impaired (≥ 3 points) (Satake, Kinoshita, Matsui, & Arai, 2020). Subjective cognitive function was assessed using a 3-point subscale of the KCL and dichotomized as not impaired (0 points) and impaired (≥ 1 point) (Tomata et al., 2017). Drinking was dichotomized as never/past and current. Smoking was dichotomized as never/past and current.

### Statistical analysis

First, the descriptive statistics of the participants were calculated. Second, the PERMA-Profiler scores at baseline and follow-up were summarized. Third, to examine the association between arts and cultural engagement and subsequent well-being, we applied a multivariable linear regression analysis and obtained unstandardized coefficients and 95% confidence intervals (CIs) for five domain scores of the PERMA-Profiler at follow-up. Three analytical models were conducted: Model 1 was adjusted for age, gender, and the corresponding PERMA-Profiler domain score at baseline; Model 2 was adjusted for all the covariates, with active and receptive engagement introduced in the analytical model separately; and Model 3 included both active and receptive engagement simultaneously and was adjusted for the same covariates as Model 2. We also performed sub-analyses on the four domains of PERMA-Profiler filler items. In the physical health domain, self-reported health was not included in the analytical model because of its conceptual overlap with other variables. Finally, to flexibly examine the frequency-response patterns between arts and cultural engagement and subsequent well-being, we fitted regression models using restricted cubic splines with four knots, treating the monthly frequency of engagement as a continuous variable.

We conducted exploratory subgroup analyses to examine whether the associations between arts and cultural engagement and subsequent well-being varied according to age group (< 75 years and ≥ 75 years), gender, educational attainment (low/middle [< 10 and 10–12 years] and high [≥ 13 years], and subjective economic status (low/middle and high). For gender, we conducted a women-only analysis because the number of male participants was small.

To address the selection bias due to dropout at follow-up, we applied inverse probability of censoring weighting (IPCW) (Seaman & White, 2013). IPCW can account for selective attrition by applying statistical weights depending on one’s probability of being followed and analyzed. Weights were calculated using the covariates, including baseline demographic, socioeconomic, and health status characteristics. Estimation of the regression analysis was performed with robust standard errors. Using sample weights, we confirmed that baseline participants’ characteristics were appropriately balanced between dropouts and follow-up (Supplementary Table 1).

To mitigate the potential bias owing to the missing values, we applied missing-value imputation using chained random forests, based on the random forest algorithm (Stekhoven & Bühlmann, 2011).

Statistical significance was set at < 0.05. All statistical analyses were conducted using R software (Version 4.5.2 for Windows; R Foundation for Statistical Computing, Vienna, Austria).

## Results

Data from 354 participants were analyzed. Table 1 shows the baseline characteristics of the participants after imputation and weighting (unweighted characteristics and missing information for each variable are shown in Supplementary Table 2). The participants’ mean age was 74.0 (standard deviation [SD] = 6.1) and 78.6% were women. Regarding arts and cultural engagement, active engagement was reported yearly to monthly by 11.8% and monthly or more by 63.2%, whereas 25.0% reported no engagement. Receptive engagement was reported yearly to monthly by 56.0% and monthly or more by 20.7%, whereas 23.3% reported no engagement.

**Table 1.**
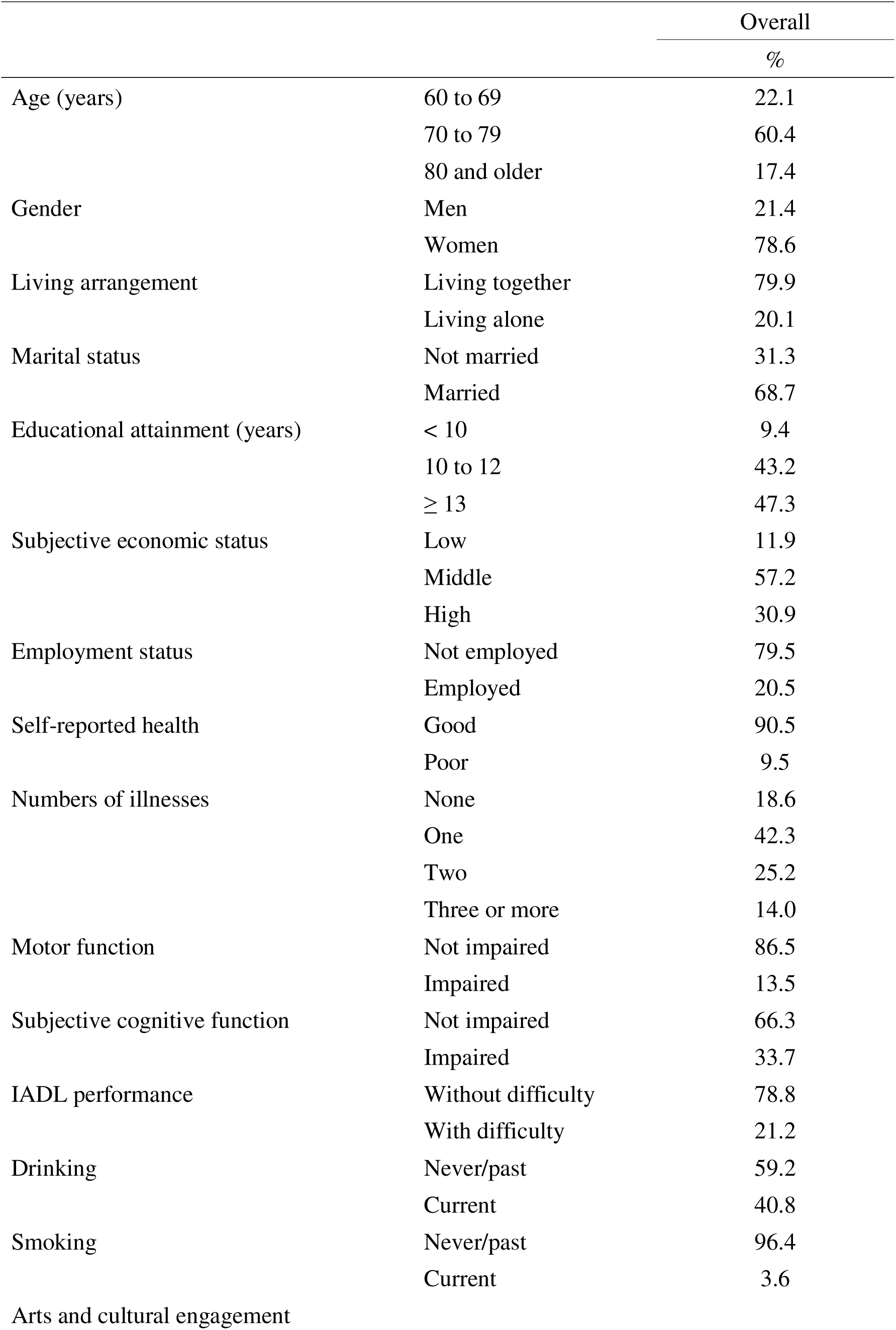
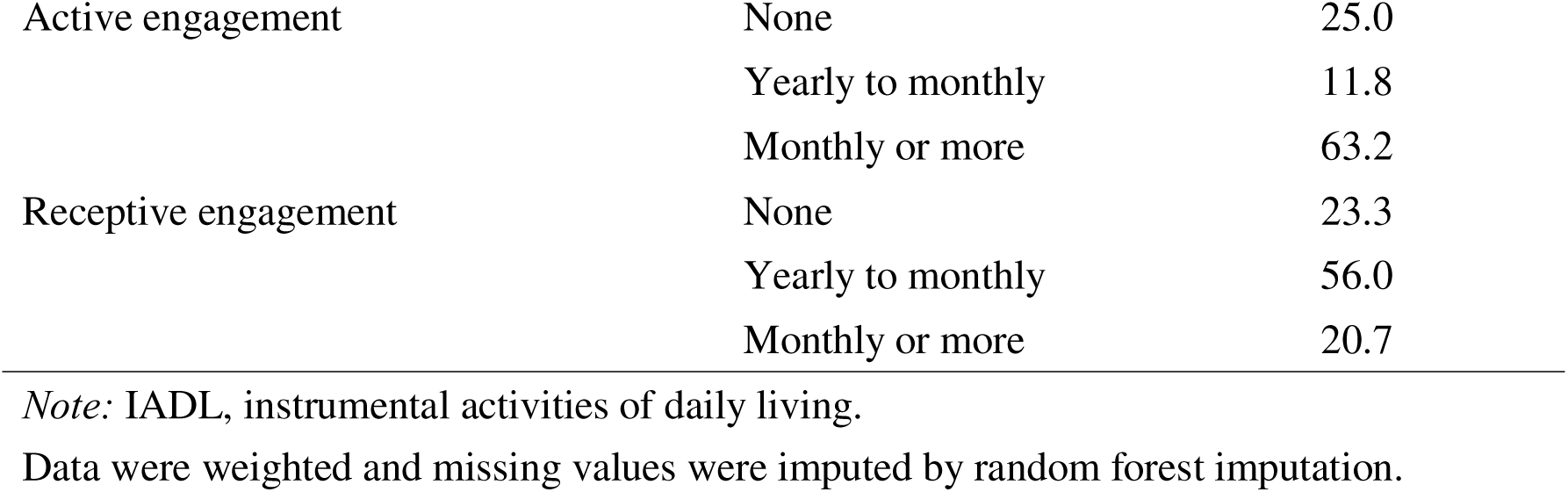
The characteristics of the participants at baseline (n = 354)

|  |  | Overall |
| --- | --- | --- |
|  |  | % |
| Age (years) | 60 to 69 | 22.1 |
|  | 70 to 79 | 60.4 |
|  | 80 and older | 17.4 |
| Gender | Men | 21.4 |
|  | Women | 78.6 |
| Living arrangement | Living together | 79.9 |
|  | Living alone | 20.1 |
| Marital status | Not married | 31.3 |
|  | Married | 68.7 |
| Educational attainment (years) | < 10 | 9.4 |
|  | 10 to 12 | 43.2 |
|  | ≥ 13 | 47.3 |
| Subjective economic status | Low | 11.9 |
|  | Middle | 57.2 |
|  | High | 30.9 |
| Employment status | Not employed | 79.5 |
|  | Employed | 20.5 |
| Self-reported health | Good | 90.5 |
|  | Poor | 9.5 |
| Numbers of illnesses | None | 18.6 |
|  | One | 42.3 |
|  | Two | 25.2 |
|  | Three or more | 14.0 |
| Motor function | Not impaired | 86.5 |
|  | Impaired | 13.5 |
| Subjective cognitive function | Not impaired | 66.3 |
|  | Impaired | 33.7 |
| IADL performance | Without difficulty | 78.8 |
|  | With difficulty | 21.2 |
| Drinking | Never/past | 59.2 |
|  | Current | 40.8 |
| Smoking | Never/past | 96.4 |
|  | Current | 3.6 |
| Arts and cultural engagement |  |  |
| Active engagement | None | 25.0 |
|  | Yearly to monthly | 11.8 |
|  | Monthly or more | 63.2 |
| Receptive engagement | None | 23.3 |
|  | Yearly to monthly | 56.0 |
|  | Monthly or more | 20.7 |

*Note:* IADL, instrumental activities of daily living.
Data were weighted and missing values were imputed by random forest imputation.

Supplementary Table 2 presents descriptive statistics of well-being at baseline and follow-up according to arts and cultural engagement. Participants with more frequent active engagement had higher scores at baseline and follow-up. Across the two time points, scores tended to decrease among participants with no engagement, whereas scores were sustained or improved among those with monthly or more frequent engagement. Similar patterns were broadly observed for receptive engagement, although differences in the domains of engagement and relationships appeared smaller than those in the other domains.

Table 2 indicates the association between arts and cultural engagement and well-being two years later. In the Model 3, multivariable linear regression analysis after adjusting for the covariates showed that, compared with non-engagement, higher frequencies of active arts and cultural engagement were positively associated with higher levels of all PERMA domains, with positive emotion (yearly to monthly: coef. = -0.21 [95% CI = -0.75, 0.32]; monthly or more: coef. = 0.62 [95% CI = 0.25, 0.99]; *P* for trend < 0.001), engagement (yearly to monthly, coef. = 0.08 [95% CI = -0.48, 0.64]; monthly or more, coef. = 0.60 [95% CI = 0.22, 0.99]; *P* for trend = 0.002), relationships (yearly to monthly, coef. = 0.09 [95% CI = -0.47, 0.65]; monthly to more, coef. = 0.74 [95% CI = 0.31, 1.17]; *P* for trend < 0.001), meaning (yearly to monthly, coef. = 0.09 [95% CI = -0.53, 0.71]; monthly or more, coef. = 0.55 [95% CI = 0.10, 1.00]; *P* for trend = 0.010), and accomplishment (yearly to monthly, coef. = 0.20 [95% CI = -0.41, 0.81]; monthly or more, coef. = 0.78 [95% CI = 0.31, 1.26]; *P* for trend < 0.001).

**Table 2.**
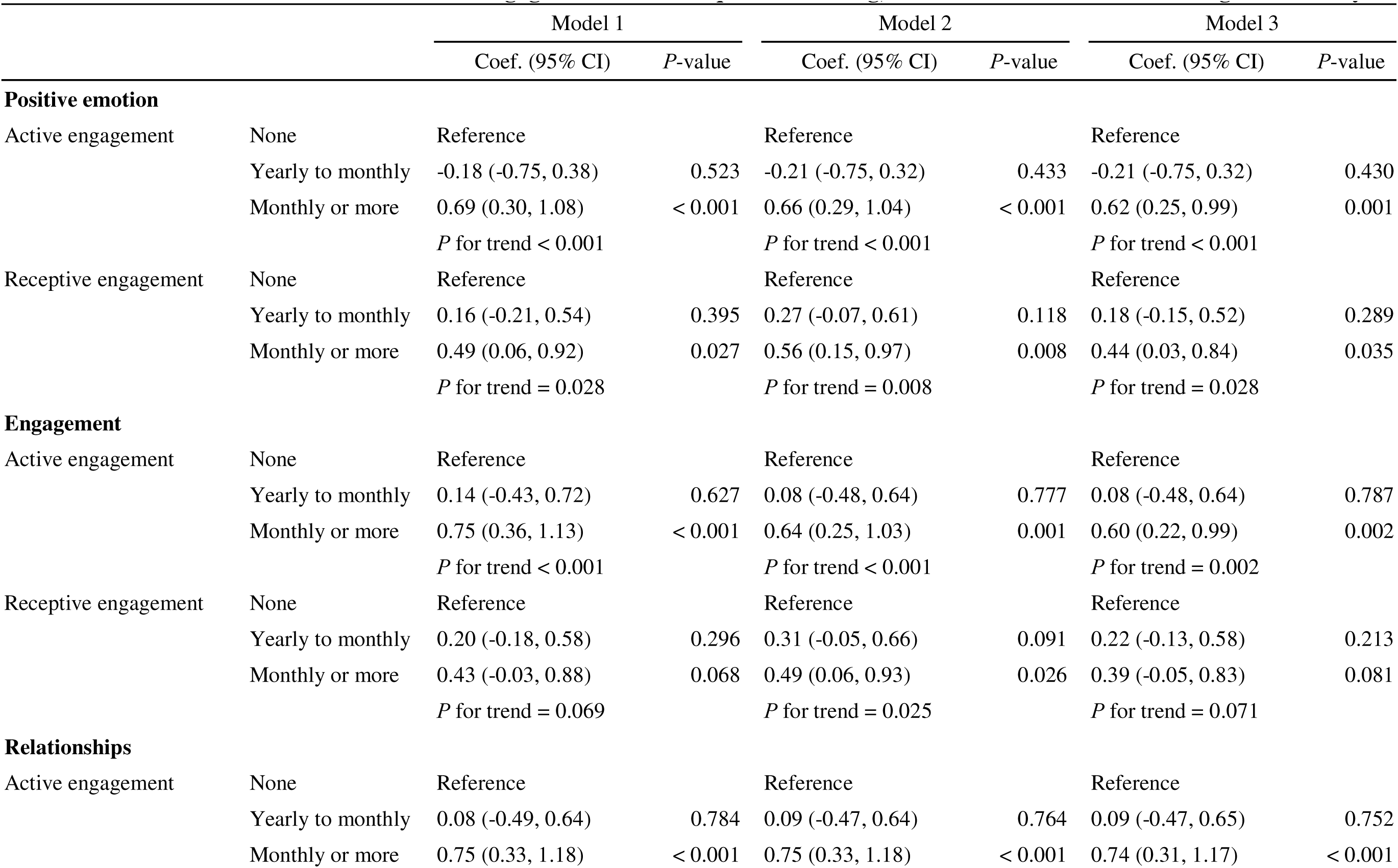
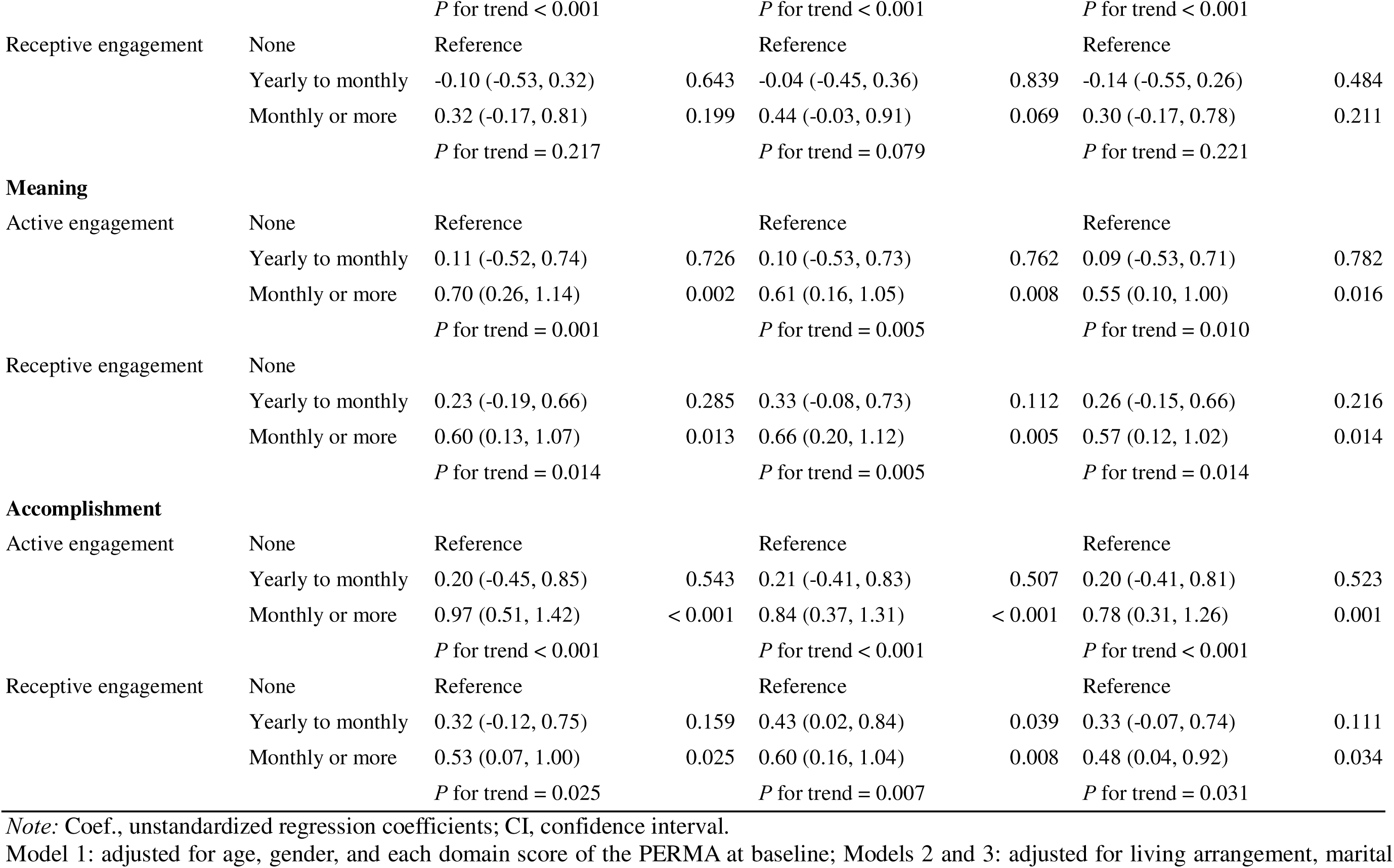
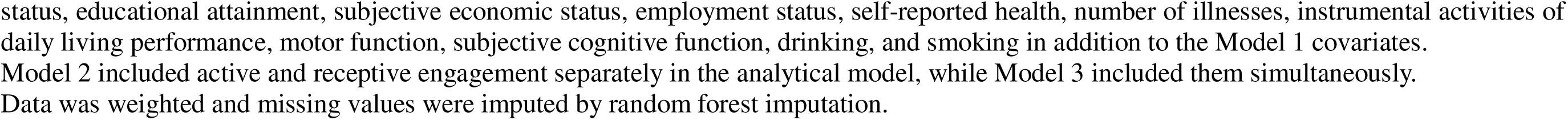
Association between arts and cultural engagement and subsequent well-being, based on multivariable linear regression analysis.

For receptive engagement, higher frequencies were positively associated with higher levels of the domains of positive emotion (Model 3 in Table 3: yearly to monthly, coef. = 0.18 [95% CI = -0.15, 0.52]; monthly or more, coef. = 0.44 [95% CI = 0.03, 0.84]; *P* for trend = 0.028), meaning (yearly to monthly, coef. = 0.26 [95% CI = -0.15, 0.66]; monthly or more, coef. = 0.57 [95% CI = 0.12, 1.02]; *P* for trend = 0.014), and accomplishment (yearly to monthly, coef. = 0.33 [95% CI = -0.07, 0.74]; monthly or more: coef. = 0.48 [95% CI = 0.04, 0.92]; *P* for trend = 0.031). In contrast, receptive engagement was not clearly associated with the domains of engagement (yearly to monthly, coef. = 0.22 [95% CI = -0.13, 0.58]; monthly or more: coef. = 0.39 [95% CI = -0.05, 0.83]; *P* for trend = 0.071) and relationships (yearly to monthly, coef. = -0.14 [95% CI = -0.55, 0.26]; monthly or more, coef. = 0.30 [95% CI = -0.17, 0.78]; *P* for trend = 0.221).

Figures 2 and 3 present the restricted cubic spline analyses that suggested frequency-response associations between the frequencies of active and receptive arts and cultural engagement and subsequent well-being, respectively. For active engagement, PERMA scores generally increased with greater frequencies, with positive emotion, engagement, relationships, and accomplishment showing steeper increases at the level of multiple monthly engagements, followed by a plateau at higher levels. Meaning score showed a more gradual positive association across the range of engagement. For receptive engagement, the scores increased at levels of engagement less than a month but tended to plateau or slightly decrease at higher levels for several domains; estimate intervals were wider at higher levels of engagement, suggesting greater uncertainty in this range.

**Figure 2.**
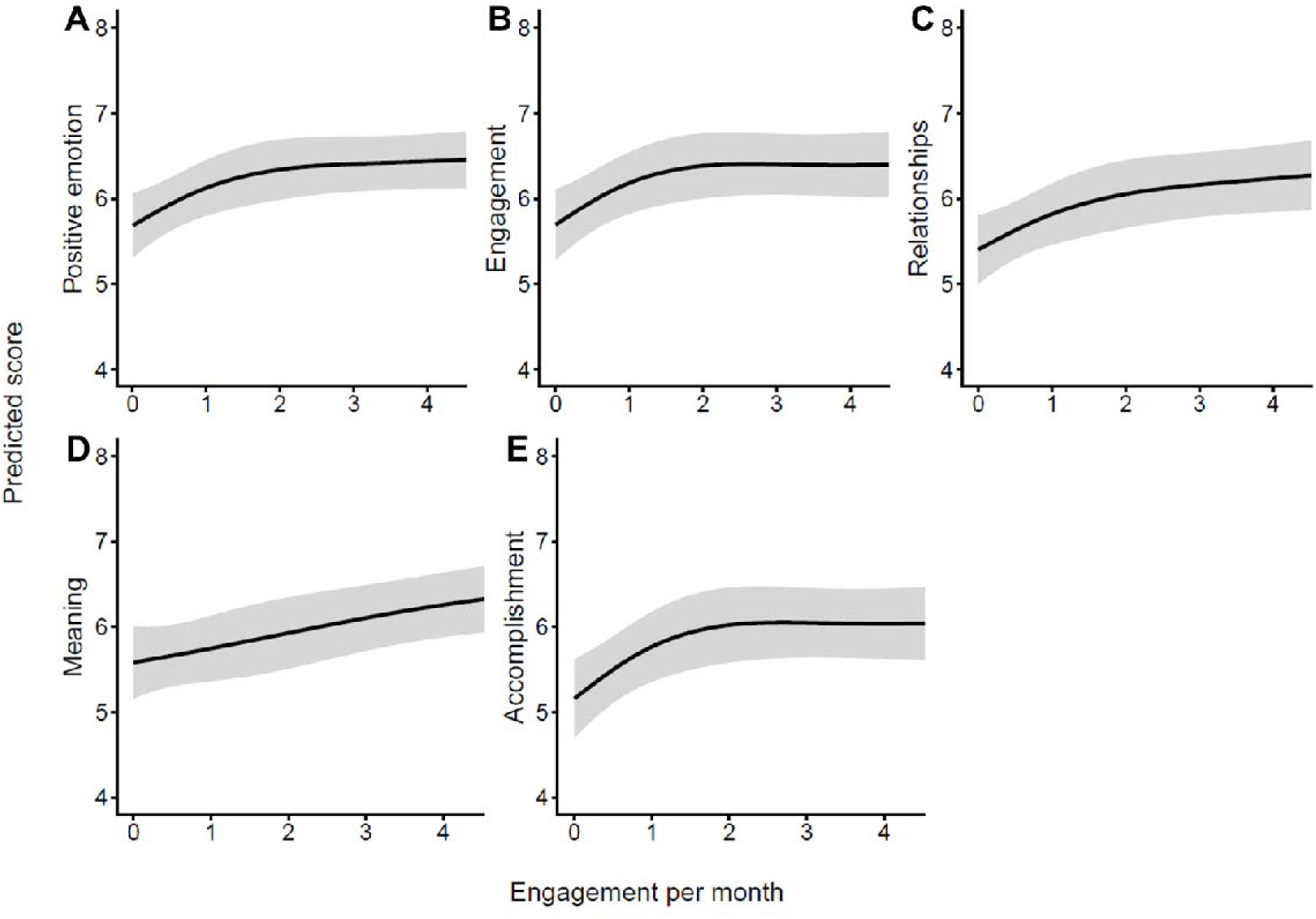
Restricted cubic spline curves for the association between active arts and cultural engagement and subsequent well-being. The curves show the adjusted predicted scores for five PERMA domains two years later: (A) positive emotion, (B) engagement, (C) relationships, (D) meaning, and (E) accomplishment. The horizontal axis indicates the frequency of active arts and cultural engagement per month, and the vertical axis indicates the predicted score. Solid black lines represent adjusted predicted values, and gray shaded areas represent 95% confidence intervals. Models were weighted with stabilized inverse probability for censoring and robust variance was used for estimation.

**Figure 3.**
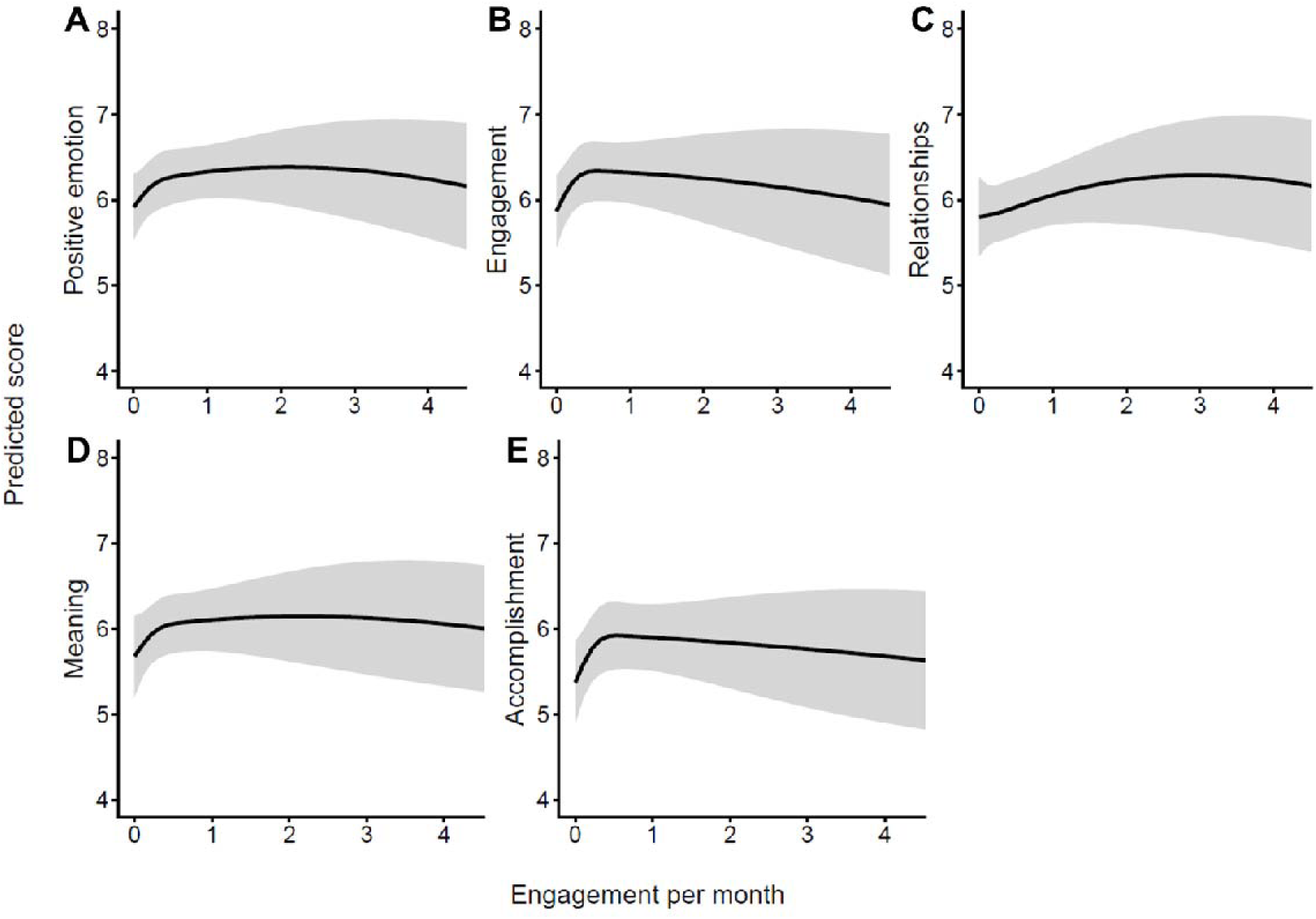
Restricted cubic spline curves for the association between receptive arts and cultural engagement and subsequent well-being. The curves show the adjusted predicted scores for five PERMA domains two years later: (A) positive emotion, (B) engagement, (C) relationships, (D) meaning, and (E) accomplishment. The horizontal axis indicates the frequency of receptive arts and cultural engagement per month, and the vertical axis indicates the predicted score. Solid black lines represent adjusted predicted values, and gray shaded areas represent 95% confidence intervals. Models were weighted with stabilized inverse probability for censoring and robust variance was used for estimation.

In the exploratory subgroup analyses (Supplementary Tables 4 to 7), the associations between arts and cultural engagement and subsequent well-being were generally consistent with the main findings, although the strength and consistency of the associations varied across subgroups. In the age-subgroup analysis, the associations were more evident among participants aged 75 years and older. The women-only analysis showed patterns broadly consistent with the main analysis. In the subgroup analyses by educational attainment and economic status, positive associations were observed in several domains, particularly among participants with low or middle educational attainment or economic status.

Supplementary Tables 8 and 9 show the descriptive statistics and regression analyses for the PERMA filler domains, respectively. After adjusting for the covariates, active engagement was associated with higher overall well-being and physical health and lower loneliness, while receptive engagement was associated with higher overall well-being and lower negative emotion. These frequency-response associations showed broadly consistent patterns, with predicted scores generally improving at higher levels of engagement, although estimation intervals widened at higher frequencies (Supplementary Figures 1 and 2).

## Discussion

This longitudinal study examined the association between arts and cultural engagement, both in the active and receptive forms, and subsequent multidimensional well-being among older adults in Japan. Our results indicated that engaging in arts and cultural activities was linked to higher levels of well-being, including multidimensional aspects, two years later. These findings suggest that arts and culture are modifiable social and community determinants of supporting well-being in later life, highlighting the importance of ensuring access to arts and culture for all as social and cultural resources.

Previous longitudinal studies of older adults in Western countries have shown that engaging in arts and culture may contribute to enhancing well-being, including hedonic and eudaimonic aspects (Bone, et al., 2022; Davies, et al., 2023; Fancourt & Tymoszuk, 2019; Finn, et al., 2025; Tymoszuk, Perkins, Spiro, et al., 2020). Although there are slight differences in the specific aspects of well-being that benefit across studies, participation in arts and culture overall appears to have positive influences on multiple areas of well-being. These support the plausibility of the present study using the PERMA framework to measure well-being and extend the evidence on potential common benefits across different cultural contexts in Western countries and East Asia. In Asian countries, including Japan, opportunities to engage with arts and culture are generally fewer than in Western countries (Agency for Cultural Affairs, 2021; Naoe, 2003), and there are some differences in their values and closeness to life. These may reflect national and traditional contexts of arts and culture, including educational opportunities, resource availability, national investment levels, and types and forms of engagements and activities. Despite these differences, arts and culture may have implications for multidimensional well-being in later life across countries and cultures.

The spline analyses suggested frequency-response patterns between arts and cultural engagement and subsequent well-being. For active engagement, well-being scores generally increased with greater frequency of engagement, although the increases appeared to level off at higher frequencies (i.e., multiple times per month). This pattern suggests that regular active engagement may be associated with better well-being, although the incremental gains may become smaller at higher frequencies. Meanwhile, for receptive engagement, the curves suggested increases in well-being at lower to moderate levels of engagement (i.e., a few times per year), followed by a plateau or slight decrease at higher levels in several domains.

Although the confidence intervals were wider at higher frequencies, with greater uncertainty in these estimates, this pattern suggests that higher frequencies of receptive engagement may not necessarily be associated with progressively higher well-being. Receptive engagement may support well-being not only through frequent attendance but also through aesthetic experiences, emotional stimulation, and a sense of connection to cultural life. From a social identity perspective (Haslam, Jetten, Cruwys, Dingle, & Haslam, 2018), even occasional engagement may help older adults perceive themselves as members of arts and culture-related communities or as people who participate in meaningful cultural life. This sense of membership might support belonging and positive self-concept. These findings should be interpreted as exploratory visual evidence that complements the regression analyses rather than as definitive evidence of a threshold or causal dose-response relationship.

Activities related to arts and culture involve various and complex components, such as aesthetics, imagination, evocation of emotion, and social interaction (Fancourt & Finn, 2019). These diverse active ingredients can contribute to people’s health and well-being through psychological, social, physical, and behavioral mechanisms, including fulfilling psychological needs, strengthening social connections, reducing stress responses, and promoting health-related behaviors (Fancourt, et al., 2021). Arts and culture also provide opportunities for creation, practice, role-taking, and interaction with others. These experiences may support well-being including meaning in life and self-fulfillment by enabling older adults to develop skills and participate in purposeful activities. Meanwhile, the findings on PERMA filler items may provide some of these interpretations. Active forms may improve physical health through physical activity and cognitive tasks related to activities and participation, and they may support the fulfillment of social relationships through shared activities, which may contribute to well-being. Meanwhile, receptive forms, which involves exposure to imaginative and extraordinary environments such as cultural institutions and events, may enhance well-being through emotional regulations, including reducing negative feelings. Although this study did not fully capture the diverse influences of arts and culture and could not formally examine mediating mechanisms, these findings highlight the necessity of further investigations to identify the detailed mechanisms linking arts and cultural engagement to well-being.

Regarding forms of engagement, active engagement showed positive associations with all domains, while receptive engagement showed associations with the domains of positive emotion, meaning, and accomplishment, but only marginal associations with engagement and relationships. Arts and cultural engagement can be divided into active forms, in which individuals participate as “an actor,” and receptive forms, in which as “an audience.” Although these share some ingredients, each form is presumed to contain specific active ingredients that may improve well-being. Prior research has suggested that receptive forms may involve lower experiential engagement, such as immersion and enthusiasm, than active forms. This interpretation is consistent with our findings (Davies, et al., 2012; Liu, et al., 2026). Engagement domain refers, for instance, to how an individual’s attention is attracted, drawn, and held by something with positive feelings, such as playfulness or enjoyment (Reschly, Huebner, Appleton, & Antaramian, 2008). Receptive forms do not necessarily indicate “passivity,” but may activate this aspect modestly. Meanwhile, some previous studies have suggested the contribution of receptive engagement to social well-being, such as reducing loneliness and obtaining social satisfaction (Tymoszuk, Perkins, Fancourt, & Williamon, 2020), which somewhat contrasts with our findings. This inconsistency may be attributed to differences in cultures and attitudes toward arts and cultural activities across countries. In the Japanese context, some receptive activities related to arts and culture, such as museum and gallery visits, may be undertaken alone or independently (Cross Marketing Inc., 2024). Such modes of engagement may not always involve sustained social interaction, which may partly explain the weaker associations with social relationship satisfaction. Nonetheless, given that our results showed a tendency toward positive associations, although the effect sizes were not large, further investigations with larger samples are warranted.

The exploratory subgroup analyses suggested possible heterogeneity for arts and cultural engagement on well-being. Notably, positive associations between engaging in arts and culture and well-being were broadly relevant among participants of older age and those with lower socioeconomic status, including lower education and economic status. These findings suggest that arts and cultural engagement may be particularly relevant for relatively vulnerable populations. Old-old adults often face social and functional losses due to bereavement, loss of friends, and age-related functional decline. For these individuals, arts and culture may serve as social assets that provide opportunities to maintain social connections and participate in communities. Additionally, given that social disparities in a range of resources by socioeconomic status are often reported in mental health issues (Fancourt & Warran, 2024; Mak, Coulter, & Fancourt, 2021), arts and culture may serve as an equitable and implementable approach to supporting individuals’ well-being, including older adults(Rodriguez, et al., 2024). Taken together, arts and culture may not be merely “leisure” activities but may also have the potential to function as public health assets for addressing inequalities related to social and functional vulnerability in aging or socioeconomic disadvantage.

There are several limitations in this study. First, because of the observational nature of the study, we could not determine the causal effects of arts and cultural engagement on well-being. Unmeasured confounders, such as life-course factors and personality traits, may have led to an overestimation of the associations. Future intervention studies and natural experiments are needed to clarify causal relationships. Second, information obtained from self-administered questionnaires may have caused measurement bias. Particularly, the frequency of arts and cultural engagement may have been misclassified. Third, the follow-up rate was moderate (67.8%), which may have resulted in selection bias. Although we partially addressed this issue using weighting for censoring, residual bias may remain. Fourth, the follow-up period was limited to two years, and the long-term associations between arts and cultural engagement and well-being remain unclear. Future studies with longer follow-up are needed. Finally, the sample size in this study was modest, and the study was based on a convenience sample recruited from public facility users. A disproportionate sample of healthy and active individuals may have caused selection bias. Furthermore, the participants were recruited from one area of Japan. The representativeness of this sample was therefore limited, restricting the generalizability of the results. Future investigations using larger and more representative data are warranted.

Despite the above limitations, the findings suggest a beneficial role of arts and culture in older adults’ well-being, not only in Western countries but also in East Asia, despite cultural differences. With the global rollout of social prescribing, referrals to arts and cultural activities are positioned as one of the key elements (Bu et al., 2025). Additionally, in the United States and the United Kingdom, there are several policies and projects that link the arts with people’s health and well-being (All-Party Parliamentary Group on Arts Health and Wellbeing, 2017; Dow, Warran, Letrondo, & Fancourt, 2023; Rodriguez et al., 2025). In Japan, there are no specific policies on arts and health or well-being, but social prescribing schemes linking arts and culture are also attracting attention. Still, access to arts and culture is limited for some groups, such as those with socioeconomic disadvantages, reflecting societal disparities. Given that arts and culture can be considered public health assets, this study emphasizes the importance of promoting them to enhance the well-being of older adults.

## Conclusions

This study suggests that arts and cultural engagement, in both active and receptive forms, may contribute to multifaceted aspects of well-being among older adults in Japan. These findings highlight the importance of ensuring accessible and inclusive arts and cultural opportunities as social and cultural resources for supporting participation, well-being, and flourishing in later life.

## Acknowledgements

We wish to express our sincere gratitude to Rieko Kato and the staff of Ai-Ai Mind Inc., for their contributions to this study. We also thank all the participants and corporators of this study and Prof. Toru Shiotani, Kanazawa Institute of Technology.

## Data availability

The data that support the findings of this study are available on request from the corresponding author. The data are not publicly available because of privacy and ethical restrictions.

## Author contributions

TN conceptualized and designed the study, participated in the data collection, analyzed the data, and drafted and revised the manuscript. ES was involved in the data collection and reviewed and critically revised the manuscript. TH reviewed and critically revised the manuscript. All authors approved the submission of the final manuscript.

