## Supplementary Tables and Figures for "Arts and Cultural Engagement and Multidimensional Well-being in Later Life"

**Supplementary Table 1. The baseline characteristics of dropout and follow-up participants before and after weighting**

|  |  | Before weighting | | | |  | After weighting | | | |
| --- | --- | --- | --- | --- | --- | --- | --- | --- | --- | --- |
|  |  | Dropout | Follow-up | *P*-value | SMD |  | Dropout | Follow-up | *P*-value | SMD |
| Age (years), % | 60 to 69 | 30.8 | 19.5 | 0.008 | 0.288 |  | 24.1 | 22.1 | 0.879 | 0.050 |
|  | 70 to 79 | 46.7 | 59.3 |  |  |  | 59.5 | 60.4 |  |  |
|  | 80 and older | 22.5 | 21.2 |  |  |  | 16.3 | 17.4 |  |  |
| Gender, % | Women | 79.3 | 77.7 | 0.762 | 0.039 |  | 79.5 | 78.6 | 0.810 | 0.024 |
| Living arrangement, % | Living alone | 23.1 | 16.7 | 0.102 | 0.161 |  | 20.0 | 20.1 | 0.965 | 0.004 |
| Marital status, % | Married | 65.7 | 72.6 | 0.109 | 0.179 |  | 67.8 | 68.7 | 0.851 | 0.019 |
| Educational attainment (years), % | < 10 | 10.7 | 11.0 | 0.658 | 0.125 |  | 8.4 | 9.4 | 0.790 | 0.068 |
|  | 10 to 12 | 42.0 | 46.3 |  |  |  | 41.0 | 43.2 |  |  |
|  | ≥ 13 | 47.3 | 42.4 |  |  |  | 50.6 | 47.3 |  |  |
| Subjective economic status, % | Low | 10.7 | 8.2 | 0.605 | 0.092 |  | 11.6 | 11.9 | 0.971 | 0.024 |
|  | Middle | 53.3 | 56.5 |  |  |  | 58.4 | 57.2 |  |  |
|  | High | 36.1 | 35.3 |  |  |  | 30.0 | 30.9 |  |  |
| Employment status, % | Employed | 21.9 | 20.1 | 0.655 | 0.082 |  | 20.5 | 20.5 | 0.996 | 0.001 |
| Self-reported health, % | Poor | 13.6 | 7.3 | 0.069 | 0.207 |  | 9.1 | 9.5 | 0.876 | 0.014 |
| Numbers of illnesses, % | None | 21.9 | 20.1 | 0.580 | 0.159 |  | 18.9 | 18.6 | 0.988 | 0.037 |
|  | One | 30.2 | 37.0 |  |  |  | 43.5 | 42.3 |  |  |
|  | Two | 24.9 | 23.7 |  |  |  | 24.6 | 25.2 |  |  |
|  | Three or more | 17.8 | 15.5 |  |  |  | 12.9 | 14.0 |  |  |
| Motor function, % | Impaired | 14.8 | 10.2 | 0.301 | 0.141 |  | 12.7 | 13.5 | 0.798 | 0.024 |
| Subjective cognitive function, % | Impaired | 39.6 | 31.6 | 0.094 | 0.209 |  | 34.8 | 33.7 | 0.821 | 0.023 |
| IADL performance, % | With difficulty | 22.5 | 20.9 | 0.556 | 0.106 |  | 19.5 | 21.2 | 0.672 | 0.042 |
| Drinking, % | Current | 41.4 | 43.2 | 0.436 | 0.138 |  | 38.9 | 40.8 | 0.718 | 0.038 |
| Smoking, % | Current | 5.3 | 2.3 | 0.114 | 0.193 |  | 3.8 | 3.6 | 0.924 | 0.010 |
| Arts and cultural engagement, % |  |  |  |  |  |  |  |  |  |  |
| Active engagement | None | 29.6 | 22.0 | 0.060 | 0.255 |  | 24.0 | 25.0 | 0.952 | 0.031 |
|  | Yearly to monthly | 10.7 | 10.2 |  |  |  | 12.6 | 11.8 |  |  |
|  | Monthly or more | 45.0 | 57.1 |  |  |  | 63.4 | 63.2 |  |  |
| Receptive engagement | None | 25.4 | 23.2 | 0.077 | 0.254 |  | 21.7 | 23.3 | 0.867 | 0.055 |
|  | Yearly to monthly | 59.2 | 51.4 |  |  |  | 55.6 | 56.0 |  |  |
|  | Monthly or more | 13.6 | 21.8 |  |  |  | 22.7 | 20.7 |  |  |
| PERMA-Profiler scores, mean (SD) |  |  |  |  |  |  |  |  |  |  |
| Positive emotion |  | 6.3 (1.8) | 6.6 (1.9) | 0.105 | 0.157 |  | 6.4 (1.8) | 6.5 (1.9) | 0.785 | 0.028 |
| Engagement |  | 6.2 (1.9) | 6.4 (1.9) | 0.144 | 0.140 |  | 6.4 (1.8) | 6.4 (1.9) | 0.956 | 0.006 |
| Relationships |  | 5.4 (1.9) | 5.7 (2.0) | 0.073 | 0.175 |  | 5.6 (1.8) | 5.7 (2.0) | 0.751 | 0.032 |
| Meaning |  | 5.9 (1.8) | 6.0 (2.1) | 0.493 | 0.068 |  | 6.0 (2.0) | 6.0 (2.1) | 0.891 | 0.014 |
| Accomplishment |  | 5.7 (1.8) | 5.9 (1.9) | 0.218 | 0.120 |  | 5.8 (1.9) | 5.8 (1.8) | 0.791 | 0.027 |
| Overall well-being |  | 6.0 (1.6) | 6.2 (1.7) | 0.153 | 0.145 |  | 6.0 (1.6) | 6.1 (1.7) | 0.700 | 0.038 |
| Physical health |  | 6.3 (2.0) | 6.7 (2.0) | 0.019 | 0.222 |  | 6.6 (2.0) | 6.5 (2.0) | 0.616 | 0.050 |
| Negative emotion |  | 3.0 (1.9) | 2.8 (1.8) | 0.346 | 0.089 |  | 3.0 (2.0) | 3.0 (1.9) | 0.812 | 0.025 |
| Loneliness |  | 2.7 (2.4) | 2.6 (2.5) | 0.644 | 0.044 |  | 2.7 (2.4) | 2.6 (2.3) | 0.643 | 0.050 |

*Note:* IADL, instrumental activities of daily living; SD, standard deviation; SMD, standard mean difference.

**Supplementary Table 2. The baseline characteristics of the participants without imputation and weighting (n = 354)**

|  |  | Overall |
| --- | --- | --- |
|  |  | % |
| Age (years) | 60 to 69 | 69 ( 19.5) |
|  | 70 to 79 | 210 ( 59.3) |
|  | 80 and older | 75 ( 21.2) |
| Gender | Men | 79 ( 22.3) |
|  | Women | 275 ( 77.7) |
| Living arrangement | Living together | 295 ( 83.3) |
|  | Living alone | 59 ( 16.7) |
| Marital status | Not married | 97 ( 27.4) |
|  | Married | 257 ( 72.6) |
| Educational attainment (years) | < 10 | 39 ( 11.0) |
|  | 10 to 12 | 164 ( 46.3) |
|  | ≥ 13 | 150 ( 42.4) |
|  | Missing | 1 ( 0.3) |
| Subjective economic status | Low | 29 ( 8.2) |
|  | Middle | 200 ( 56.5) |
|  | High | 125 ( 35.3) |
| Employment status | Not employed | 281 ( 79.4) |
|  | Employed | 71 ( 20.1) |
|  | Missing | 2 ( 0.6) |
| Self-reported health | Good | 316 ( 89.3) |
|  | Poor | 26 ( 7.3) |
|  | Missing | 12 ( 3.4) |
| Numbers of illnesses | None | 71 ( 20.1) |
|  | One | 131 ( 37.0) |
|  | Two | 84 ( 23.7) |
|  | Three or more | 55 ( 15.5) |
|  | Missing | 13 ( 3.7) |
| Motor function | Not impaired | 254 ( 71.8) |
|  | Impaired | 36 ( 10.2) |
|  | Missing | 64 ( 18.1) |
| Subjective cognitive function | Not impaired | 227 ( 64.1) |
|  | Impaired | 112 ( 31.6) |
|  | Missing | 15 ( 4.2) |
| IADL performance | Without difficulty | 268 ( 75.7) |
|  | With difficulty | 74 ( 20.9) |
|  | Missing | 12 ( 3.4) |
| Drinking | Never/past | 198 ( 55.9) |
|  | Current | 153 ( 43.2) |
|  | Missing | 3 ( 0.8) |
| Smoking | Never/past | 344 ( 97.2) |
|  | Current | 8 ( 2.3) |
|  | Missing | 2 ( 0.6) |
| Arts and cultural engagement |  |  |
| Active engagement | None | 78 ( 22.0) |
|  | Yearly to monthly | 36 ( 10.2) |
|  | Monthly or more | 202 ( 57.1) |
|  | Missing | 38 ( 10.7) |
| Receptive engagement | None | 82 ( 23.2) |
|  | Yearly to monthly | 182 ( 51.4) |
|  | Monthly or more | 77 ( 21.8) |
|  | Missing | 13 ( 3.7) |
| PERMA-Profiler scores, mean (SD)* |  |  |
| Positive emotion |  | 6.6 (1.9) |
| Engagement |  | 6.4 (1.9) |
| Relationships |  | 5.7 (2.0) |
| Meaning |  | 6.0 (2.1) |
| Accomplishment |  | 5.9 (1.9) |
| Overall well-being |  | 6.2 (1.7) |
| Physical health |  | 6.7 (2.0) |
| Negative emotion |  | 2.8 (1.8) |
| Loneliness |  | 2.6 (2.5) |

*Note:* IADL, instrumental activities of daily living; SD, standard deviation.

*Missing values: positive emotion, n = 24; engagement, n = 28; relationships, n = 33; meaning, n = 36; accomplishment, n = 25; overall well-being, n = 69; physical health, n = 17; negative emotion, n = 19; loneliness, n = 14.

**Supplementary Table 3. PERMA-Profiler scores at baseline and follow-up according to arts and cultural engagement**

|  | Positive emotion | |  | Engagement | |  | Relationships | |  | Meaning | |  | Accomplishment | |
| --- | --- | --- | --- | --- | --- | --- | --- | --- | --- | --- | --- | --- | --- | --- |
|  | Baseline | Follow-up |  | Baseline | Follow-up |  | Baseline | Follow-up |  | Baseline | Follow-up |  | Baseline | Follow-up |
|  | Mean (SD) | Mean (SD) |  | Mean (SD) | Mean (SD) |  | Mean (SD) | Mean (SD) |  | Mean (SD) | Mean (SD) |  | Mean (SD) | Mean (SD) |
| Active engagement |  |  |  |  |  |  |  |  |  |  |  |  |  |  |
| None | 5.6 (2.0) | 5.3 (2.2) |  | 5.6 (1.9) | 5.4 (2.1) |  | 5.2 (2.2) | 5.0 (1.9) |  | 5.3 (2.1) | 5.2 (2.2) |  | 5.1 (2.0) | 4.9 (2.1) |
| Yearly to monthly | 6.5 (1.6) | 5.7 (1.9) |  | 6.5 (1.5) | 6.1 (1.9) |  | 5.6 (1.7) | 5.3 (1.9) |  | 5.8 (2.1) | 5.7 (1.9) |  | 5.7 (2.0) | 5.5 (2.2) |
| Monthly or more | 6.8 (1.7) | 6.8 (1.8) |  | 6.7 (1.8) | 6.8 (1.8) |  | 5.9 (1.9) | 6.7 (1.8) |  | 6.4 (1.9) | 6.6 (1.9) |  | 6.2 (1.7) | 6.5 (1.8) |
| Receptive engagement |  |  |  |  |  |  |  |  |  |  |  |  |  |  |
| None | 6.0 (2.0) | 5.7 (2.0) |  | 6.0 (1.9) | 5.9 (2.0) |  | 5.2 (1.9) | 5.5 (2.0) |  | 5.6 (2.3) | 5.6 (2.3) |  | 5.5 (1.9) | 5.4 (1.9) |
| Yearly to monthly | 6.5 (1.8) | 6.3 (2.0) |  | 6.3 (1.8) | 6.3 (2.0) |  | 5.7 (1.9) | 5.7 (1.8) |  | 6.1 (2.0) | 6.1 (2.0) |  | 5.8 (1.8) | 6.0 (2.1) |
| Monthly or more | 7.0 (1.8) | 7.0 (1.9) |  | 6.9 (1.8) | 6.9 (1.8) |  | 6.1 (2.2) | 6.3 (1.9) |  | 6.4 (2.0) | 6.7 (1.9) |  | 6.3 (1.8) | 6.5 (1.9) |

*Note:* SD, standard deviation.

Data were weighted and missing values were imputed by random forest imputation.

**Supplementary Table 4. Subgroup analysis by age on association between arts and cultural engagement and subsequent well-being, based on multivariable linear regression analysis**

|  |  | Under 75 years old | |  | Aged 75 and older | |
| --- | --- | --- | --- | --- | --- | --- |
|  |  | Coef. (95% CI) | *P*-value |  | Coef. (95% CI) | *P*-value |
| **Positive emotion** |  |  |  |  |  |  |
| Active engagement | None | Reference |  |  | Reference |  |
|  | Yearly to monthly | -0.78 (-1.54, -0.01) | 0.048 |  | 0.25 (-0.46, 0.97) | 0.491 |
|  | Monthly or more | 0.33 (-0.14, 0.81) | 0.172 |  | 0.91 (0.42, 1.40) | < 0.001 |
|  |  | *P* for trend = 0.075 |  |  | *P* for trend < 0.001 |  |
| Receptive engagement | None | Reference |  |  | Reference |  |
|  | Yearly to monthly | -0.16 (-0.57, 0.26) | 0.460 |  | 0.49 (-0.01, 1.00) | 0.059 |
|  | Monthly or more | 0.25 (-0.29, 0.79) | 0.370 |  | 0.76 (0.19, 1.34) | 0.011 |
|  |  | *P* for trend = 0.431 |  |  | *P* for trend = 0.007 |  |
| **Engagement** |  |  |  |  |  |  |
| Active engagement | None | Reference |  |  | Reference |  |
|  | Yearly to monthly | -0.29 (-1.19, 0.61) | 0.525 |  | 0.41 (-0.28, 1.11) | 0.241 |
|  | Monthly or more | 0.36 (-0.17, 0.90) | 0.182 |  | 0.86 (0.36, 1.37) | 0.001 |
|  |  | *P* for trend = 0.133 |  |  | *P* for trend < 0.001 |  |
| Receptive engagement | None | Reference |  |  | Reference |  |
|  | Yearly to monthly | -0.19 (-0.70, 0.31) | 0.459 |  | 0.62 (0.09, 1.14) | 0.023 |
|  | Monthly or more | 0.05 (-0.56, 0.67) | 0.861 |  | 0.69 (0.04, 1.33) | 0.039 |
|  |  | *P* for trend = 0.890 |  |  | *P* for trend = 0.025 |  |
| **Relationships** |  |  |  |  |  |  |
| Active engagement | None | Reference |  |  | Reference |  |
|  | Yearly to monthly | -0.69 (-1.47, 0.10) | 0.090 |  | 0.96 (0.16, 1.75) | 0.019 |
|  | Monthly or more | 0.50 (-0.05, 1.05) | 0.074 |  | 1.12 (0.55, 1.70) | < 0.001 |
|  |  | *P* for trend = 0.041 |  |  | *P* for trend < 0.001 |  |
| Receptive engagement | None | Reference |  |  | Reference |  |
|  | Yearly to monthly | -0.38 (-0.98, 0.21) | 0.210 |  | 0.06 (-0.52, 0.64) | 0.836 |
|  | Monthly or more | 0.14 (-0.52, 0.81) | 0.669 |  | 0.70 (0.03, 1.37) | 0.044 |
|  |  | *P* for trend = 0.732 |  |  | *P* for trend = 0.083 |  |
| **Meaning** |  |  |  |  |  |  |
| Active engagement | None | Reference |  |  | Reference |  |
|  | Yearly to monthly | 0.00 (-0.80, 0.80) | 0.994 |  | 0.36 (-0.44, 1.17) | 0.379 |
|  | Monthly or more | 0.56 (-0.04, 1.16) | 0.068 |  | 0.64 (0.05, 1.24) | 0.036 |
|  |  | *P* for trend = 0.051 |  |  | *P* for trend = 0.026 |  |
| Receptive engagement | None | Reference |  |  | Reference |  |
|  | Yearly to monthly | -0.10 (-0.67, 0.47) | 0.729 |  | 0.71 (0.12, 1.29) | 0.020 |
|  | Monthly or more | 0.37 (-0.28, 1.02) | 0.269 |  | 0.88 (0.22, 1.54) | 0.010 |
|  |  | *P* for trend = 0.280 |  |  | *P* for trend = 0.009 |  |
| **Accomplishment** |  |  |  |  |  |  |
| Active engagement | None | Reference |  |  | Reference |  |
|  | Yearly to monthly | 0.23 (-0.58, 1.04) | 0.585 |  | 0.27 (-0.57, 1.11) | 0.537 |
|  | Monthly or more | 0.54 (-0.07, 1.15) | 0.084 |  | 1.14 (0.48, 1.79) | 0.001 |
|  |  | *P* for trend = 0.073 |  |  | *P* for trend < 0.001 |  |
| Receptive engagement | None | Reference |  |  | Reference |  |
|  | Yearly to monthly | 0.00 (-0.53, 0.53) | 0.991 |  | 0.80 (0.18, 1.42) | 0.013 |
|  | Monthly or more | 0.12 (-0.43, 0.67) | 0.664 |  | 0.80 (0.09, 1.50) | 0.028 |
|  |  | *P* for trend = 0.666 |  |  | *P* for trend = 0.016 |  |

*Note:* Coef., unstandardized regression coefficients; CI, confidence interval.

Adjusted for gender, living arrangement, marital status, educational attainment, subjective economic status, employment status, self-reported health, number of illnesses, instrumental activities of daily living performance, motor function, subjective cognitive function, drinking smoking, and each domain score of the PERMA at baseline. Active and receptive engagement were simultaneously included in the analytical model.

Data were weighted and missing values were imputed by random forest imputation.

**Supplementary Table 5. Subgroup analysis of women on association between arts and cultural engagement and subsequent well-being, based on multivariable linear regression analysis**

|  |  | Women | |
| --- | --- | --- | --- |
|  |  | Coef. (95% CI) | *P*-value |
| **Positive emotion** |  |  |  |
| Active engagement | None | Reference |  |
|  | Yearly to monthly | -0.04 (-0.78, 0.69) | 0.907 |
|  | Monthly or more | 0.58 (0.15, 1.00) | 0.009 |
|  |  | *P* for trend = 0.004 |  |
| Receptive engagement | None | Reference |  |
|  | Yearly to monthly | 0.35 (-0.04, 0.74) | 0.083 |
|  | Monthly or more | 0.43 (-0.06, 0.93) | 0.087 |
|  |  | *P* for trend = 0.070 |  |
| **Engagement** |  |  |  |
| Active engagement | None | Reference |  |
|  | Yearly to monthly | 0.00 (-0.77, 0.77) | 0.995 |
|  | Monthly or more | 0.62 (0.17, 1.08) | 0.007 |
|  |  | *P* for trend = 0.003 |  |
| Receptive engagement | None | Reference |  |
|  | Yearly to monthly | 0.40 (-0.01, 0.81) | 0.055 |
|  | Monthly or more | 0.32 (-0.21, 0.85) | 0.241 |
|  |  | *P* for trend = 0.203 |  |
| **Relationships** |  |  |  |
| Active engagement | None | Reference |  |
|  | Yearly to monthly | 0.16 (-0.57, 0.89) | 0.665 |
|  | Monthly or more | 0.58 (0.09, 1.08) | 0.022 |
|  |  | *P* for trend = 0.022 |  |
| Receptive engagement | None | Reference |  |
|  | Yearly to monthly | -0.07 (-0.55, 0.4) | 0.759 |
|  | Monthly or more | 0.37 (-0.18, 0.92) | 0.188 |
|  |  | *P* for trend = 0.179 |  |
| **Meaning** |  |  |  |
| Active engagement | None | Reference |  |
|  | Yearly to monthly | 0.02 (-0.74, 0.77) | 0.965 |
|  | Monthly or more | 0.48 (-0.03, 0.99) | 0.064 |
|  |  | *P* for trend = 0.040 |  |
| Receptive engagement | None | Reference |  |
|  | Yearly to monthly | 0.51 (0.03, 0.99) | 0.036 |
|  | Monthly or more | 0.72 (0.17, 1.26) | 0.010 |
|  |  | *P* for trend = 0.009 |  |
| **Accomplishment** |  |  |  |
| Active engagement | None | Reference |  |
|  | Yearly to monthly | 0.010 (-0.76, 0.78) | 0.978 |
|  | Monthly or more | 0.72 (0.14, 1.29) | 0.015 |
|  |  | *P* for trend = 0.009 |  |
| Receptive engagement | None | Reference |  |
|  | Yearly to monthly | 0.46 (-0.05, 0.96) | 0.080 |
|  | Monthly or more | 0.57 (0.01, 1.12) | 0.046 |
|  |  | *P* for trend = 0.040 |  |

*Note:* Coef., unstandardized regression coefficients; CI, confidence interval.

Adjusted for age, living arrangement, marital status, educational attainment, subjective economic status, employment status, self-reported health, number of illnesses, instrumental activities of daily living performance, motor function, subjective cognitive function, drinking, smoking, and each domain score of the PERMA at baseline. Active and receptive engagement were simultaneously included in the analytical model.

Data were weighted and missing values were imputed by random forest imputation.

**Supplementary Table 6. Subgroup analysis by educational attainment on association between arts and cultural engagement and subsequent well-being, based on multivariable linear regression analysis**

|  |  | Low/middle | |  | High | |
| --- | --- | --- | --- | --- | --- | --- |
|  |  | Coef. (95% CI) | *P*-value |  | Coef. (95% CI) | *P*-value |
| **Positive emotion** |  |  |  |  |  |  |
| Active engagement | None | Reference |  |  | Reference |  |
|  | Yearly to monthly | -0.70 (-1.41, 0.01) | 0.054 |  | 0.08 (-0.71, 0.86) | 0.848 |
|  | Monthly or more | 0.89 (0.45, 1.32) | < 0.001 |  | 0.43 (-0.14, 1.00) | 0.144 |
|  |  | *P* for trend < 0.001 |  |  | *P* for trend = 0.125 |  |
| Receptive engagement | None | Reference |  |  | Reference |  |
|  | Yearly to monthly | 0.38 (-0.04, 0.80) | 0.077 |  | 0.23 (-0.31, 0.78) | 0.405 |
|  | Monthly or more | 0.55 (0.01, 1.08) | 0.047 |  | 0.43 (-0.16, 1.02) | 0.155 |
|  |  | *P* for trend = 0.051 |  |  | *P* for trend = 0.145 |  |
| **Engagement** |  |  |  |  |  |  |
| Active engagement | None | Reference |  |  | Reference |  |
|  | Yearly to monthly | -0.73 (-1.46, 0.00) | 0.052 |  | 0.59 (-0.21, 1.39) | 0.151 |
|  | Monthly or more | 0.58 (0.11, 1.05) | 0.016 |  | 0.68 (0.11, 1.26) | 0.021 |
|  |  | *P* for trend = 0.006 |  |  | *P* for trend = 0.020 |  |
| Receptive engagement | None | Reference |  |  | Reference |  |
|  | Yearly to monthly | 0.33 (-0.11, 0.77) | 0.148 |  | 0.35 (-0.23, 0.92) | 0.239 |
|  | Monthly or more | 0.62 (0.11, 1.14) | 0.018 |  | 0.08 (-0.59, 0.75) | 0.817 |
|  |  | *P* for trend = 0.027 |  |  | *P* for trend = 0.816 |  |
| **Relationships** |  |  |  |  |  |  |
| Active engagement | None | Reference |  |  | Reference |  |
|  | Yearly to monthly | -0.08 (-0.92, 0.76) | 0.854 |  | 0.19 (-0.54, 0.92) | 0.619 |
|  | Monthly or more | 0.79 (0.21, 1.38) | 0.009 |  | 0.54 (-0.05, 1.14) | 0.075 |
|  |  | *P* for trend = 0.007 |  |  | *P* for trend = 0.076 |  |
| Receptive engagement | None | Reference |  |  | Reference |  |
|  | Yearly to monthly | 0.16 (-0.40, 0.73) | 0.574 |  | -0.20 (-0.78, 0.37) | 0.488 |
|  | Monthly or more | 0.61 (-0.08, 1.30) | 0.084 |  | 0.14 (-0.53, 0.81) | 0.689 |
|  |  | *P* for trend = 0.094 |  |  | *P* for trend = 0.712 |  |
| **Meaning** |  |  |  |  |  |  |
| Active engagement | None | Reference |  |  | Reference |  |
|  | Yearly to monthly | 0.26 (-0.54, 1.06) | 0.527 |  | -0.05 (-0.88, 0.77) | 0.896 |
|  | Monthly or more | 0.90 (0.35, 1.45) | 0.002 |  | 0.27 (-0.39, 0.93) | 0.419 |
|  |  | *P* for trend = 0.001 |  |  | *P* for trend = 0.370 |  |
| Receptive engagement | None | Reference |  |  | Reference |  |
|  | Yearly to monthly | 0.11 (-0.4, 0.62) | 0.676 |  | 0.52 (-0.11, 1.16) | 0.110 |
|  | Monthly or more | 0.57 (0.02, 1.12) | 0.046 |  | 0.62 (-0.11, 1.36) | 0.099 |
|  |  | *P* for trend = 0.046 |  |  | *P* for trend = 0.086 |  |
| **Accomplishment** |  |  |  |  |  |  |
| Active engagement | None | Reference |  |  | Reference |  |
|  | Yearly to monthly | 0.01 (-0.76, 0.79) | 0.976 |  | 0.18 (-0.72, 1.07) | 0.701 |
|  | Monthly or more | 1.31 (0.62, 1.99) | < 0.001 |  | 0.38 (-0.28, 1.04) | 0.261 |
|  |  | *P* for trend < 0.001 |  |  | *P* for trend = 0.237 |  |
| Receptive engagement | None | Reference |  |  | Reference |  |
|  | Yearly to monthly | 0.34 (-0.23, 0.92) | 0.245 |  | 0.55 (-0.03, 1.14) | 0.067 |
|  | Monthly or more | 0.59 (-0.04, 1.21) | 0.070 |  | 0.46 (-0.19, 1.11) | 0.167 |
|  |  | *P* for trend = 0.078 |  |  | *P* for trend = 0.157 |  |

*Note:* Coef., unstandardized regression coefficients; CI, confidence interval.

Adjusted for age, gender, living arrangement, marital status, subjective economic status, employment status, self-reported health, number of illnesses, instrumental activities of daily living performance, motor function, subjective cognitive function, drinking, smoking, and each domain score of the PERMA at baseline. Active and receptive engagement were simultaneously included in the analytical model.

Data were weighted and missing values were imputed by random forest imputation.

**Supplementary Table 7. Subgroup analysis by economic status on association between arts and cultural engagement and subsequent well-being, based on multivariable linear regression analysis**

|  |  | Low/middle | |  | High | |
| --- | --- | --- | --- | --- | --- | --- |
|  |  | Coef. (95% CI) | *P*-value |  | Coef. (95% CI) | *P*-value |
| **Positive emotion** |  |  |  |  |  |  |
| Active engagement | None | Reference |  |  | Reference |  |
|  | Yearly to monthly | -0.15 (-0.78, 0.48) | 0.648 |  | 0.24 (-0.73, 1.22) | 0.624 |
|  | Monthly or more | 0.63 (0.21, 1.05) | 0.004 |  | 0.74 (0.13, 1.36) | 0.021 |
|  |  | *P* for trend = 0.002 |  |  | *P* for trend = 0.012 |  |
| Receptive engagement | None | Reference |  |  | Reference |  |
|  | Yearly to monthly | 0.18 (-0.25, 0.60) | 0.416 |  | 0.23 (-0.32, 0.77) | 0.413 |
|  | Monthly or more | 0.56 (0.04, 1.08) | 0.036 |  | 0.47 (-0.11, 1.04) | 0.115 |
|  |  | *P* for trend = 0.036 |  |  | *P* for trend = 0.090 |  |
| **Engagement** |  |  |  |  |  |  |
| Active engagement | None | Reference |  |  | Reference |  |
|  | Yearly to monthly | 0.27 (-0.39, 0.92) | 0.422 |  | 0.32 (-0.52, 1.16) | 0.455 |
|  | Monthly or more | 0.85 (0.41, 1.29) | < 0.001 |  | 0.07 (-0.59, 0.72) | 0.844 |
|  |  | *P* for trend < 0.001 |  |  | *P* for trend = 0.815 |  |
| Receptive engagement | None | Reference |  |  | Reference |  |
|  | Yearly to monthly | 0.24 (-0.20, 0.69) | 0.288 |  | 0.35 (-0.16, 0.85) | 0.186 |
|  | Monthly or more | 0.46 (-0.12, 1.04) | 0.124 |  | 0.44 (-0.21, 1.09) | 0.191 |
|  |  | *P* for trend = 0.121 |  |  | *P* for trend = 0.239 |  |
| **Relationships** |  |  |  |  |  |  |
| Active engagement | None | Reference |  |  | Reference |  |
|  | Yearly to monthly | 0.00 (-0.67, 0.66) | 0.989 |  | 0.84 (-0.11, 1.79) | 0.086 |
|  | Monthly or more | 0.76 (0.25, 1.27) | 0.004 |  | 0.72 (-0.04, 1.49) | 0.067 |
|  |  | *P* for trend = 0.002 |  |  | *P* for trend = 0.151 |  |
| Receptive engagement | None | Reference |  |  | Reference |  |
|  | Yearly to monthly | 0.06 (-0.45, 0.58) | 0.811 |  | -0.09 (-0.73, 0.55) | 0.778 |
|  | Monthly or more | 0.38 (-0.24, 1.01) | 0.232 |  | 0.57 (-0.17, 1.31) | 0.133 |
|  |  | *P* for trend = 0.243 |  |  | *P* for trend = 0.157 |  |
| **Meaning** |  |  |  |  |  |  |
| Active engagement | None | Reference |  |  | Reference |  |
|  | Yearly to monthly | 0.09 (-0.70, 0.88) | 0.826 |  | 0.36 (-0.68, 1.40) | 0.499 |
|  | Monthly or more | 0.64 (0.14, 1.14) | 0.013 |  | 0.51 (-0.30, 1.31) | 0.221 |
|  |  | *P* for trend = 0.008 |  |  | *P* for trend = 0.235 |  |
| Receptive engagement | None | Reference |  |  | Reference |  |
|  | Yearly to monthly | 0.38 (-0.13, 0.89) | 0.149 |  | 0.10 (-0.55, 0.75) | 0.766 |
|  | Monthly or more | 0.77 (0.17, 1.36) | 0.012 |  | 0.53 (-0.22, 1.27) | 0.168 |
|  |  | *P* for trend = 0.014 |  |  | *P* for trend = 0.153 |  |
| **Accomplishment** |  |  |  |  |  |  |
| Active engagement | None | Reference |  |  | Reference |  |
|  | Yearly to monthly | 0.25 (-0.55, 1.05) | 0.540 |  | 0.59 (-0.52, 1.70) | 0.298 |
|  | Monthly or more | 0.95 (0.40, 1.49) | < 0.001 |  | 0.31 (-0.52, 1.14) | 0.469 |
|  |  | *P* for trend < 0.001 |  |  | *P* for trend = 0.655 |  |
| Receptive engagement | None | Reference |  |  | Reference |  |
|  | Yearly to monthly | 0.45 (-0.03, 0.94) | 0.069 |  | -0.06 (-0.8, 0.69) | 0.880 |
|  | Monthly or more | 0.66 (0.10, 1.21) | 0.022 |  | 0.37 (-0.41, 1.16) | 0.357 |
|  |  | *P* for trend = 0.022 |  |  | *P* for trend = 0.389 |  |

*Note:* Coef., unstandardized regression coefficients; CI, confidence interval.

Adjusted for age, gender, living arrangement, marital status, educational attainment, employment status, self-reported health, number of illnesses, instrumental activities of daily living performance, motor function, subjective cognitive function, drinking, smoking, and each domain score of the PERMA at baseline. Active and receptive engagement were simultaneously included in the analytical model.

Data were weighted and missing values were imputed by random forest imputation.

**Supplementary Table 8. PERMA-Profiler filler domain scores at baseline and follow-up according to arts and cultural engagement**

|  | Overall well-being | |  | Physical health | |  | Negative emotion | |  | Loneliness | |
| --- | --- | --- | --- | --- | --- | --- | --- | --- | --- | --- | --- |
|  | Baseline | Follow-up |  | Baseline | Follow-up |  | Baseline | Follow-up |  | Baseline | Follow-up |
|  | Mean (SD) | Mean (SD) |  | Mean (SD) | Mean (SD) |  | Mean (SD) | Mean (SD) |  | Mean (SD) | Mean (SD) |
| Active engagement |  |  |  |  |  |  |  |  |  |  |  |
| None | 5.6 (2.0) | 5.3 (2.2) |  | 6.0 (2.3) | 5.6 (2.3) |  | 3.4 (1.9) | 3.8 (1.9) |  | 2.8 (2.1) | 3.8 (2.7) |
| Yearly to monthly | 6.5 (1.6) | 5.7 (1.9) |  | 6.3 (2.0) | 5.9 (2.2) |  | 3.2 (1.4) | 3.7 (1.7) |  | 2.9 (2.1) | 2.9 (2.4) |
| Monthly or more | 6.8 (1.7) | 6.8 (1.8) |  | 6.8 (1.8) | 6.8 (1.8) |  | 2.8 (1.9) | 3.1 (1.9) |  | 2.4 (2.4) | 2.6 (2.2) |
| Receptive engagement |  |  |  |  |  |  |  |  |  |  |  |
| None | 5.7 (1.8) | 5.7 (1.8) |  | 6.1 (1.8) | 6.0 (2.2) |  | 3.2 (1.7) | 3.6 (1.9) |  | 2.7 (2.2) | 3.0 (2.7) |
| Yearly to monthly | 6.2 (1.6) | 6.2 (1.7) |  | 6.6 (1.9) | 6.4 (2.0) |  | 2.9 (1.8) | 3.4 (1.9) |  | 2.5 (2.3) | 3.0 (2.3) |
| Monthly or more | 6.5 (1.8) | 6.8 (1.7) |  | 6.8 (2.1) | 6.7 (2.0) |  | 3.1 (2.2) | 3.1 (2.0) |  | 2.6 (2.5) | 2.8 (2.5) |

*Note:* SD, standard deviation.

Data were weighted and missing values were imputed by random forest imputation.

**Supplementary Table 9. Association between arts and cultural engagement and subsequent PERMA filler domain scores, based on multivariable linear regression analysis**

|  |  | Model 1 | |  | Model 2 | |  | Model 3 | |
| --- | --- | --- | --- | --- | --- | --- | --- | --- | --- |
|  |  | Coef. (95% CI) | *P*-value |  | Coef. (95% CI) | *P*-value |  | Coef. (95% CI) | *P*-value |
| **Overall well-being** |  |  |  |  |  |  |  |  |  |
| Active engagement | None | Reference |  |  | Reference |  |  | Reference |  |
|  | Yearly to monthly | 0.13 (-0.23, 0.48) | 0.481 |  | 0.12 (-0.23, 0.47) | 0.492 |  | 0.12 (-0.23, 0.47) | 0.502 |
|  | Monthly or more | 0.69 (0.40, 0.97) | < 0.001 |  | 0.69 (0.40, 0.97) | < 0.001 |  | 0.65 (0.36, 0.93) | < 0.001 |
|  |  | *P* for trend < 0.001 |  |  | *P* for trend < 0.001 |  |  | *P* for trend < 0.001 |  |
| Receptive engagement | None | Reference |  |  | Reference |  |  | Reference |  |
|  | Yearly to monthly | 0.19 (-0.11, 0.48) | 0.222 |  | 0.25 (-0.03, 0.54) | 0.078 |  | 0.17 (-0.10, 0.44) | 0.215 |
|  | Monthly or more | 0.49 (0.16, 0.81) | 0.004 |  | 0.51 (0.20, 0.83) | 0.002 |  | 0.41 (0.10, 0.72) | 0.010 |
|  |  | *P* for trend = 0.004 |  |  | *P* for trend = 0.001 |  |  | *P* for trend = 0.010 |  |
| **Physical health** |  |  |  |  |  |  |  |  |  |
| Active engagement | None | Reference |  |  | Reference |  |  | Reference |  |
|  | Yearly to monthly | 0.02 (-0.58, 0.63) | 0.945 |  | 0.03 (-0.60, 0.65) | 0.932 |  | 0.02 (-0.60, 0.65) | 0.939 |
|  | Monthly or more | 0.63 (0.25, 1.02) | 0.002 |  | 0.52 (0.14, 0.91) | 0.009 |  | 0.51 (0.12, 0.90) | 0.011 |
|  |  | *P* for trend < 0.001 |  |  | *P* for trend = 0.016 |  |  | *P* for trend = 0.022 |  |
| Receptive engagement | None | Reference |  |  | Reference |  |  | Reference |  |
|  | Yearly to monthly | 0.06 (-0.39, 0.51) | 0.796 |  | 0.14 (-0.30, 0.57) | 0.539 |  | 0.06 (-0.36, 0.48) | 0.788 |
|  | Monthly or more | 0.13 (-0.37, 0.63) | 0.614 |  | 0.19 (-0.30, 0.69) | 0.448 |  | 0.09 (-0.39, 0.58) | 0.702 |
|  |  | *P* for trend = 0.616 |  |  | *P* for trend = 0.367 |  |  | *P* for trend = 0.562 |  |
| **Negative emotion** |  |  |  |  |  |  |  |  |  |
| Active engagement | None | Reference |  |  | Reference |  |  | Reference |  |
|  | Yearly to monthly | 0.06 (-0.49, 0.61) | 0.834 |  | 0.00 (-0.54, 0.53) | 0.993 |  | 0.00 (-0.53, 0.53) | 0.988 |
|  | Monthly or more | -0.35 (-0.77, 0.06) | 0.095 |  | -0.22 (-0.62, 0.18) | 0.283 |  | -0.18 (-0.58, 0.22) | 0.382 |
|  |  | *P* for trend = 0.070 |  |  | *P* for trend = 0.258 |  |  | *P* for trend = 0.398 |  |
| Receptive engagement | None | Reference |  |  | Reference |  |  | Reference |  |
|  | Yearly to monthly | -0.03 (-0.43, 0.38) | 0.897 |  | -0.05 (-0.45, 0.35) | 0.792 |  | -0.03 (-0.43, 0.38) | 0.897 |
|  | Monthly or more | -0.46 (-0.92, 0.00) | 0.052 |  | -0.50 (-0.95, -0.05) | 0.029 |  | -0.46 (-0.92, -0.01) | 0.047 |
|  |  | *P* for trend = 0.058 |  |  | *P* for trend = 0.033 |  |  | *P* for trend = 0.048 |  |
| **Loneliness** |  |  |  |  |  |  |  |  |  |
| Active engagement | None | Reference |  |  | Reference |  |  | Reference |  |
|  | Yearly to monthly | -0.94 (-1.78, -0.11) | 0.028 |  | -0.88 (-1.71, -0.04) | 0.041 |  | -0.88 (-1.71, -0.05) | 0.038 |
|  | Monthly or more | -1.16 (-1.74, -0.57) | < 0.001 |  | -0.99 (-1.58, -0.39) | 0.001 |  | -1.01 (-1.58, -0.43) | < 0.001 |
|  |  | *P* for trend < 0.001 |  |  | *P* for trend = 0.001 |  |  | *P* for trend = 0.001 |  |
| Receptive engagement | None | Reference |  |  | Reference |  |  | Reference |  |
|  | Yearly to monthly | 0.03 (-0.56, 0.63) | 0.909 |  | 0.12 (-0.46, 0.71) | 0.683 |  | 0.25 (-0.30, 0.80) | 0.378 |
|  | Monthly or more | -0.21 (-0.89, 0.46) | 0.535 |  | -0.14 (-0.80, 0.51) | 0.670 |  | 0.02 (-0.61, 0.65) | 0.939 |
|  |  | *P* for trend = 0.548 |  |  | *P* for trend = 0.691 |  |  | *P* for trend = 0.889 |  |

*Note:* Coef., unstandardized regression coefficients; CI, confidence interval.

Model 1: adjusted for age, gender, and each domain score of the PERMA at baseline; Models 2 and 3: adjusted for living arrangement, marital status, educational attainment, subjective economic status, employment status, self-reported health, number of illnesses, instrumental activities of daily living performance, motor function, subjective cognitive function, drinking, and smoking in addition to the Model 1 covariates. For physical health, the analysis model did not include self-reported health.

Data was weighted and missing values were imputed by random forest imputation.

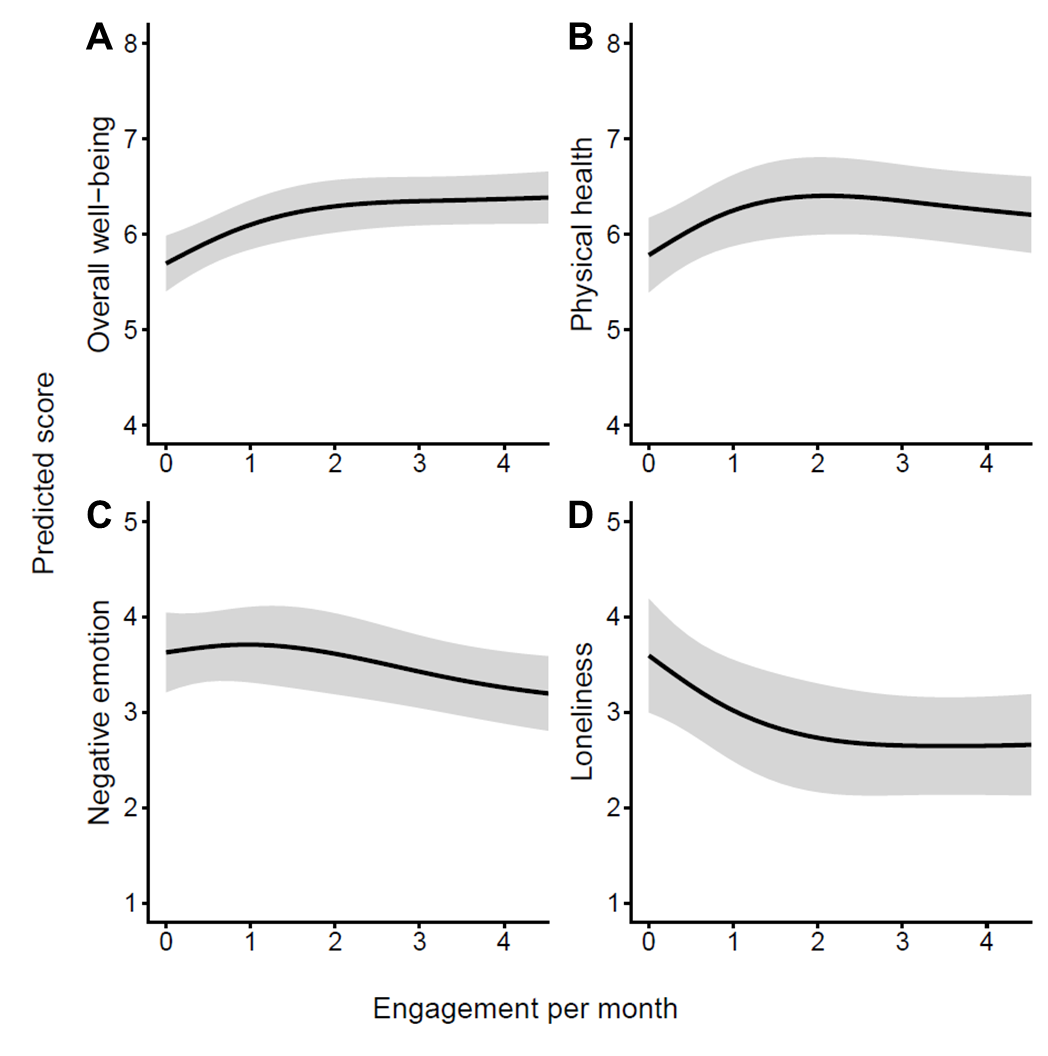

**Supplementary Figure 1. Restricted cubic spline curves for the association between active arts and cultural engagement and subsequent PERMA filler domain scores.** The curves show the adjusted predicted scores for four PERMA filler domains two years later: (A) overall well-being, (B) physical health, (C) negative emotion, and (D) loneliness. The horizontal axis indicates the frequency of active arts and cultural engagement per month, and the vertical axis indicates the predicted score. Solid black lines represent adjusted predicted values, and gray shaded areas represent 95% confidence intervals. Models were weighted with stabilized inverse probability for censoring and robust variance was used for estimation.

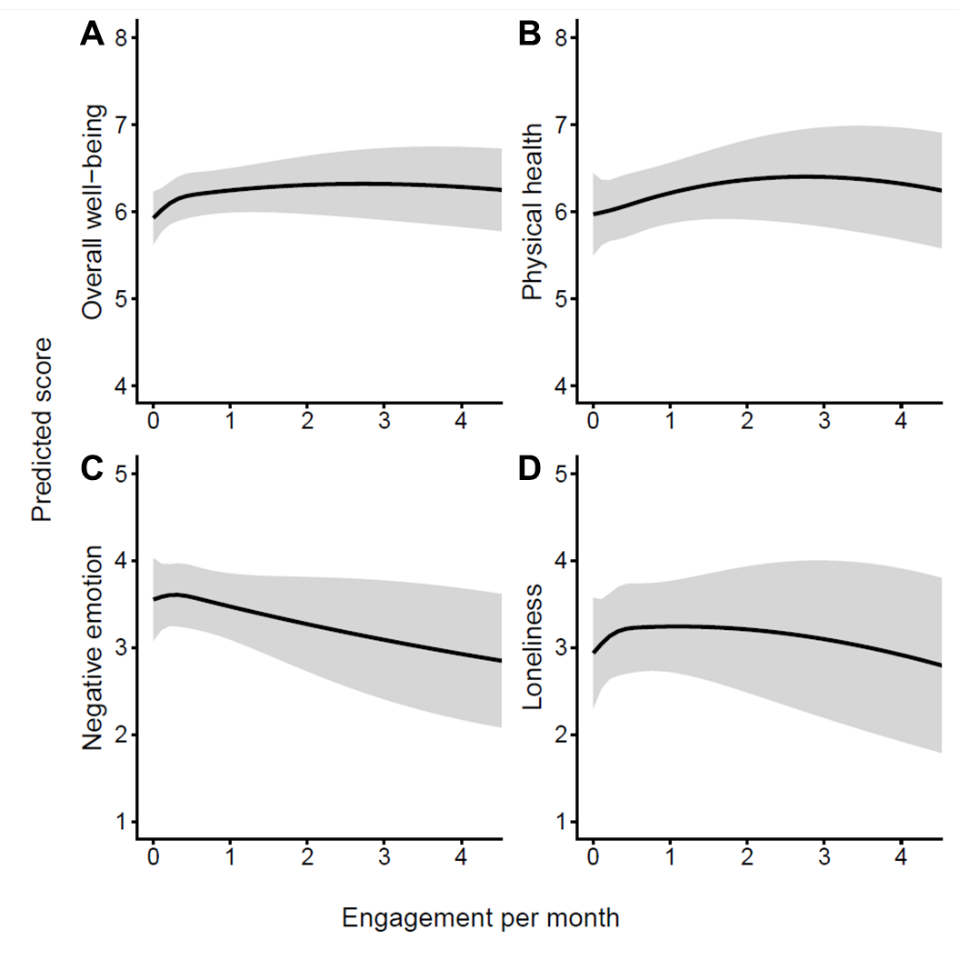

**Supplementary Figure 2. Restricted cubic spline curves for the association between receptive arts and cultural engagement and subsequent PERMA filler domain scores.** The curves show the adjusted predicted scores for four PERMA filler domains two years later: (A) overall well-being, (B) physical health, (C) negative emotion, and (D) loneliness. The horizontal axis indicates the frequency of receptive arts and cultural engagement per month, and the vertical axis indicates the predicted score. Solid black lines represent adjusted predicted values, and gray shaded areas represent 95% confidence intervals. Models were weighted with stabilized inverse probability for censoring and robust variance was used for estimation.
